# Development and external validation of prognostic models for COVID-19 to support risk stratification in secondary care

**DOI:** 10.1101/2021.01.25.21249942

**Authors:** Nicola J Adderley, Thomas Taverner, Malcolm Price, Christopher Sainsbury, David Greenwood, Joht Singh Chandan, Yemisi Takwoingi, Rashan Haniffa, Isaac Hosier, Carly Welch, Dhruv Parekh, Suzy Gallier, Krishna Gokhale, Alastair K Denniston, Elizabeth Sapey, Krishnarajah Nirantharakumar

## Abstract

**Objectives:** Existing UK prognostic models for patients admitted to hospital with COVID-19 are limited by reliance on comorbidities, which are under-recorded in secondary care, and lack of imaging data among the candidate predictors. Our aims were to develop and externally validate novel prognostic models for adverse outcomes (death, intensive therapy unit (ITU) admission) in UK secondary care; and externally validate the existing 4C score.

**Design:** Candidate predictors included demographic variables, symptoms, physiological measures, imaging, laboratory tests. Final models used logistic regression with stepwise selection.

**Setting:** Model development was performed in data from University Hospitals Birmingham (UHB). External validation was performed in the CovidCollab dataset.

**Participants:** Patients with COVID-19 admitted to UHB January-August 2020 were included.

**Main outcome measures:** Death and ITU admission within 28 days of admission.

**Results:** 1040 patients with COVID-19 were included in the derivation cohort; 288 (28%) died and 183 (18%) were admitted to ITU within 28 days of admission. Area under the receiver operating curve (AUROC) for mortality was 0.791 (95%CI 0.761-0.822) in UHB and 0.767 (95%CI 0.754-0.780) in CovidCollab; AUROC for ITU admission was 0.906 (95%CI 0.883-0.929) in UHB and 0.811 (95%CI 0.795-0.828) in CovidCollab. Models showed good calibration. Addition of comorbidities to candidate predictors did not improve model performance. AUROC for the 4C score in the UHB dataset was 0.754 (95%CI 0.721-0.786).

**Conclusions:** The novel prognostic models showed good discrimination and calibration in derivation and external validation datasets, and outperformed the existing 4C score. The models can be integrated into electronic medical records systems to calculate each individual patient’s probability of death or ITU admission at the time of hospital admission. Implementation of the models and clinical utility should be evaluated.

**Article Summary:** *Strengths and limitations of this study:*

- We developed novel prognostic models predicting mortality and ITU admission within 28 days of admission for patients hospitalised with COVID-19, using a large routinely collected dataset gathered at admission with a wide range of possible predictors (demographic variables, symptoms, physiological measures, imaging, laboratory test results).
- These novel models showed good discrimination and calibration in both derivation and external validation cohorts, and outperformed the existing ISARIC model and 4C score in the derivation dataset. We found that addition of comorbidities to the set of candidate predictors included in model derivation did not improve model performance.
- If integrated into hospital electronic medical records systems, the model algorithms will provide a predicted probability of mortality or ITU admission for each patient based on their individual data at, or close to, the time of admission, which will support clinicians’ decision making with regard to appropriate patient care pathways and triage. This information might also assist clinicians in explaining complex prognostic assessments and decisions to patients and their relatives.
- A limitation of the study was that in the external validation cohort we were unable to examine all of the predictors included in the original full UHB model due to only a reduced set of candidate predictors being available in CovidCollab. Nevertheless, the reduced model performed well and the results suggest it may be applicable in a wide range of datasets where only a reduced set of predictor variables is available.
- Furthermore, it was not possible to carry out stratified analysis by ethnicity as the UHB dataset contained too few patients in most of the strata, and no ethnicity data was available in the CovidCollab dataset.

## Background

The coronavirus disease 2019 (COVID-19) pandemic has placed exceptional strain on health care systems globally – in high income as well as low and middle income countries (LMIC). Health systems, and especially critical care services, can be overwhelmed given the number of patients, and the duration and severity of their illness. A proportion of patients with COVID-19 can deteriorate rapidly. Clinicians need to differentiate between those with COVID-19 who are at high risk of the most severe symptoms (requiring intensive care treatment/ventilation) or death, and those who can be considered low risk and potentially managed in the community. Early identification of patients at highest risk of severe outcomes may provide opportunity to prioritise, intervene and improve outcomes.

Objective prognostic tools for patients with COVID-19 would be of considerable benefit to clinicians in a secondary care setting. Prognostic models that can accurately discriminate between patients who will progress to more severe symptoms or death, and those who do not, can be used by clinicians to triage and manage patients. Such models based on patients’ initial characteristics, symptoms, biomarkers and imaging at the time of hospital admission can be used at or just after admission. This could potentially reduce time to appropriate interventions and improve patient outcomes.

A rapid systematic review has identified a number of prediction models developed for COVID-19, including prognostic models.^1^ However, while these existing studies provided useful information on candidate predictors for further exploration, there were substantial limitations: many models were developed exclusively in a Chinese population; many were at high risk of bias, particularly in terms of inclusion of non-representative control participants, inappropriate exclusion criteria, and small sample sizes leading to high risk of overfitting; most models have not considered imaging findings as candidate predictor variables; and external validation was limited.

More recent models have since been developed,^2,3^ some of which overcome a number of these limitations, including the International Severe Acute Respiratory and emerging Infection Consortium (ISARIC) model and corresponding (simplified) 4C score, which was developed in a UK secondary care population representing 260 hospitals in England, Scotland and Wales (the ISARIC dataset).^3^ Whilst the 4C score showed reasonable discrimination for mortality, there are a number of limitations, including a reliance on clinicians counting specific comorbidities, which are known to be under-recorded in secondary care,^4^ and an absence of imaging data among the candidate predictors. Furthermore, it is unclear if this model can help with predicting deteriorating patients beyond day of admission. The latter is important given the unpredictable clinical course of COVID-19 patients in the early days after admission. This raises two important research questions: first, to what extent does the inclusion of comorbidities and additional patient information (such as imaging and additional biomarkers) improve prognostic models for hospitalised patients with COVID-19? And second, is there any added value in updating the clinical parameters with evolving biomarkers to improve prediction of the clinical course of patients, with patients reassessed in real-time as the disease evolves?

The overarching aims of this study were to novel develop prognostic models for adverse outcomes (death, intensive therapy unit (ITU) admission) in a UK secondary care setting; to externally validate these models in an international dataset (including data from UK hospitals); to externally validate the existing UK ISARIC model and 4C score;^3^ and to compare performance of the newly developed models with the UK ISARIC model and 4C score. In addition, we developed daily models using time series data from the first eight days from admission to explore changes in predictors over time.

## Methods

### Data source

Data from University Hospitals Birmingham NHS Foundation Trust (UHB) was sourced via the PIONEER Health Data Research Hub for acute care, and used for model development and for external validation of the ISARIC model and 4C score. Data from patients with COVID-19 admitted to Queen Elizabeth Hospital, Birmingham (part of UHB), between 1^st^ January 2020 and 16^th^ August 2020 was included. Data included symptoms recorded at admission, comorbidities (from International Classification of Diseases 10^th^ revision (ICD-10) discharge codes), vital signs (e.g. blood pressure, oxygen saturation), laboratory results (biochemistry, haematology, microbiology, pathology), imaging, and outcomes (ITU admission and death).

External validation of the newly developed models was performed in the CovidCollab dataset. CovidCollab is an international project utilising routinely collected health care data to develop a better understanding of how best to treat and care for adults with COVID-19.^5,6^ The dataset includes symptoms, comorbidities, vital signs, laboratory results, imaging findings, and outcomes.

### Study population

Patients of all ages diagnosed with COVID-19 and hospitalised were included. Diagnosis was defined as a positive test result for SARS-CoV-2 from one or more reverse transcription polymerase chase reaction (RT-PCR) or transcription mediated amplification tests. In the CovidCollab dataset, COVID-19 diagnosis was by either PCR or antibody test.

### Study Design

Retrospective cohort analyses; index date (start of follow-up) was the hospital admission date. Study period was 1^st^ January 2020 to 12^th^ September 2020 (last admission date was 16^th^ August to ensure a minimum of 28 days of follow-up).

### Outcomes

Primary outcome was death within 28 days of admission (in-hospital or post-discharge). Secondary outcome was ITU admission within 28 days of admission.

### Study follow-up

Participants were followed from index (admission) date until the earliest of outcome date or study end (latest available data, 12^th^ September 2020). Participants were censored 28 days after index date. Participants admitted after 16^th^ August 2020 (less than 28 days prior to study end date) were excluded.

### Candidate predictor variables

Candidate predictors were selected a priori following a review of existing literature, discussion with clinical experts (specialists in acute care, critical care and geriatric medicine), and based on availability of variables routinely collected in secondary care/UHB. These included demographic variables, symptoms, comorbidities, physiological measures, imaging findings and laboratory test results. Comorbidities are not reliably and completely collected at admission, with the most complete hospital record of comorbidities usually being the discharge ICD-10 codes; therefore, the development and performance of models with and without comorbidity predictors were compared in order to explore the potential for developing models which would require no additional data collection (other than routinely collected data) at the point of admission.

### Model development

Models were trained using UHB data (patients admitted up to and including 16^th^ August 2020). We used a multi-stage model building process that assessed the impact of a range of feature representation and modelling choices to select important candidate predictors. All analyses were performed in R.

Three sets of models were fitted which incorporated continuous variables in three different ways, to explore the impact of treating these variables as continuous or categorical, and also to explore the impact of different methods of handling missing data:

a. as continuous numeric values, with missing values imputed (“continuous”);
b. as categorical values derived from the imputed continuous values (“categorical-imputed”).
c. and, in secondary analysis, as categorical values, using clinically meaningful categories and reference ranges, with missing indicators as a separate category (“categorical”);

To represent continuous variables with nonlinear relationships to the logit of the outcome, we initially fitted an exploratory predictive model (gam from the mgcv package in R), representing continuous features as thin-plate smoothed splines with the smoothing factor (gamma) set to 1.4 to shrink coefficient degrees of freedom. Based on visual inspection for obvious departure for linearity, if the variable appeared to have a linear relationship with the response term, we represented it as a linear term in variable selection and model testing and training. If the relationship between the response and the variable of interest appeared to be nonlinear, we represented the feature as a spline term (“bs” from splines package) with the minimum degrees of freedom (3 d.f.) to avoid overfitting.

For the three ways of handling numeric features and missing variables above, we fitted outcomes of death within 28 days and ITU admission (within 28 days) to candidate predictors using a range of models, which allowed both linear relationships and complex interactions between variables:

a. logistic regression with i) all baseline parameters (demographic variables, symptoms, vital signs/physiological measures, and laboratory test results), ii) demographic variables only, and iii) all baseline parameters with the addition of recorded comorbidities (recorded up to the point of discharge);
b. logistic regression with stepwise AIC minimisation, both forward and backward;^7^
c. LASSO (l1 penalized) logistic regression using all baseline parameters;
d. gradient boosted model (GBM) using all baseline parameters with default hyperparameter values of 150 trees, maximum interaction depth 3, minimum of 10 observations in nodes, and shrinkage 0.1.^8^

For each of these four variable selection models, in order to reduce overfitting and selection bias, we internally validated using 5-fold cross-validation (80/20 train/test split) to derive the candidate variable list. To avoid sensitivity to imputation, this cross-validation was repeated for each of the 5 multiple imputations.

Due to the relatively small number of outcome events (<300) we did not attempt to systematically look for interactions between multiple variables.

Model performance (discrimination) was assessed by calculating the area under the receiver operating characteristics curve (AUROC, or C-statistic).^9^ Calibration was assessed by plotting observed probability of the outcome against predicted probability, and by calculating the calibration slope and intercept. We also calculated sensitivity, specificity, positive predicted value (PPV) and negative predictive value (NPV) for the final models. For each feature set and each model, the final results for cross-validated (optimism-adjusted) AUROC and other metrics were combined from all the multiple imputations of the data set using Rubin’s rules for the mean and confidence interval (derived from the standard deviation).^10^

### Missing data

Information on candidate predictors was collected at the point of admission; however, where information on physiological or laboratory measures was not available on the day of admission, measures recorded up to 72 hours after admission were used. Candidate predictors for which >40% of patients had missing data were excluded from the analysis. Further missing continuous variables (vital signs, laboratory tests) and symptoms were imputed using the R “mice” (Multiple Imputation using Chained Equations) multiple imputation package; we performed 5 imputations, using predictive mean matching and a maximum of 50 iterations.^11^ We also explored use of a missing category for missing test results. Absence of a record of a comorbidity was taken to indicate absence of the condition.

### External validation

To investigate the transferability of models, we performed external validation of logistic regression models derived from the UHB dataset in the CovidCollab dataset, for predicting outcomes of 28-day mortality and ITU admission.

Not all candidate predictors were common to both datasets; therefore, new logistic regression models for death within 28 days and for ITU admission were re-fitted on the UHB data using only those variables also present in the CovidCollab data. We then performed an external validation of these UHB models in the CovidCollab dataset, and ascertained the AUROC in both the UHB and CovidCollab datasets. Based on model performance observed in the initial model derivation and in the interest of clinical utility, we used only categorical rather than continuous numeric variables, with imputed missing values (imputed prior to categorisation). To verify that predictors behaved similarly, we compared logistic coefficients from UHB to the same models fitted on the CovidCollab dataset. To account for sensitivity to missing values, we performed training and testing five times on 5-fold multiply imputed data sets for both UHB and CovidCollab.

### External validation of ISARIC and 4C models

An extreme gradient boosting (XGBoost) model using the ISARIC study parameters and logistic regression using the 4C score was performed in the UHB dataset (following the same modelling methods used in the original ISARIC study). The ISARIC XGBoost model was calibrated to the UHB data by deriving UHB-specific coefficients for the included variables. Model performance was assessed by calculating the AUROC and plotting calibration curves.

### Sensitivity analyses

Most patient records had some missing variables; we therefore performed a complete case analysis, where we re-fitted the best forward stepwise selection model to complete case data, then data with ≤1, 2, 5, and 10 missing values, imputing missing values in the same way as above, and examined AUROCs and logistic coefficients for stability.

In addition, we performed a sensitivity analysis within male and female strata by assessing the final model performance (AUROC) on male and female patients separately, and by comparing to performance of separate models which were fitted to male and female patients only.

### Time series analysis

The UHB regression models utilised baseline measurement data collected on admission; where not available at admission, we accepted values up to 72 hours after admission. To investigate fine-grained temporal effects of data acquisition, we produced a series of separate logistic regression models using data collected at different time windows from within 24 hours of admission up to within 7 days of admission, in 1-day increments, for the mortality outcome. Each dataset included only those patients eligible at the end of the window (not dead or discharged). This created eight different sets of predictors, including baseline variables of age, gender, symptoms, and the time-sensitive variables of the latest physiological and laboratory measurements available.

For missing data, data were carried forward from the first observation (last observation carried forward, LOCF) and 5-fold multiple imputation was performed for missing data after LOCF was done, within each separate time-window dataset. Each model was trained and tested in 5-fold cross validation, within each imputation, and AUROCs averaged using Rubin’s rule. We compared the AUROCs for each of the eight models for predicting 28-day mortality from the time of admission and compared the logistic coefficients for the models.

For additional insight into possible effects of changing measurements, we produced an additional logistic model for 28-day mortality to time-sensitive data collected within 4 days of admission, augmented with predictors indicating an increase or decrease in the category of each time-sensitive predictor relative to the reference category from 0 to 4 days – for example, whether temperature had crossed from below to above 37.8°C in that period.

### Patient and public involvement

We engaged with members of the PIONEER patient and public involvement group during development of the study protocol. We will further engage with this group, as well as other local and national patient and public involvement groups, in order to discuss dissemination of the findings and the best way to communicate these to patients and the public. We also consulted with several secondary care clinicians before and during the study to ensure that the tools developed meet the needs of clinicians. We have engaged with local NHS trusts to ensure that the algorithms developed are implemented/tested in a hospital setting.

## Results

### Derivation cohort characteristics

A total of 1040 participants with COVID-19 admitted to UHB were included in the derivation cohort. 288 (28%) died within 28 days of admission and 183 (18%) were admitted to ITU. Baseline characteristics are presented in Table 1. The mean (SD) age of participants was 68.2 (17.7) years; 57% (589) were male; and almost 90% had at least one comorbidity.

**Table 1.**
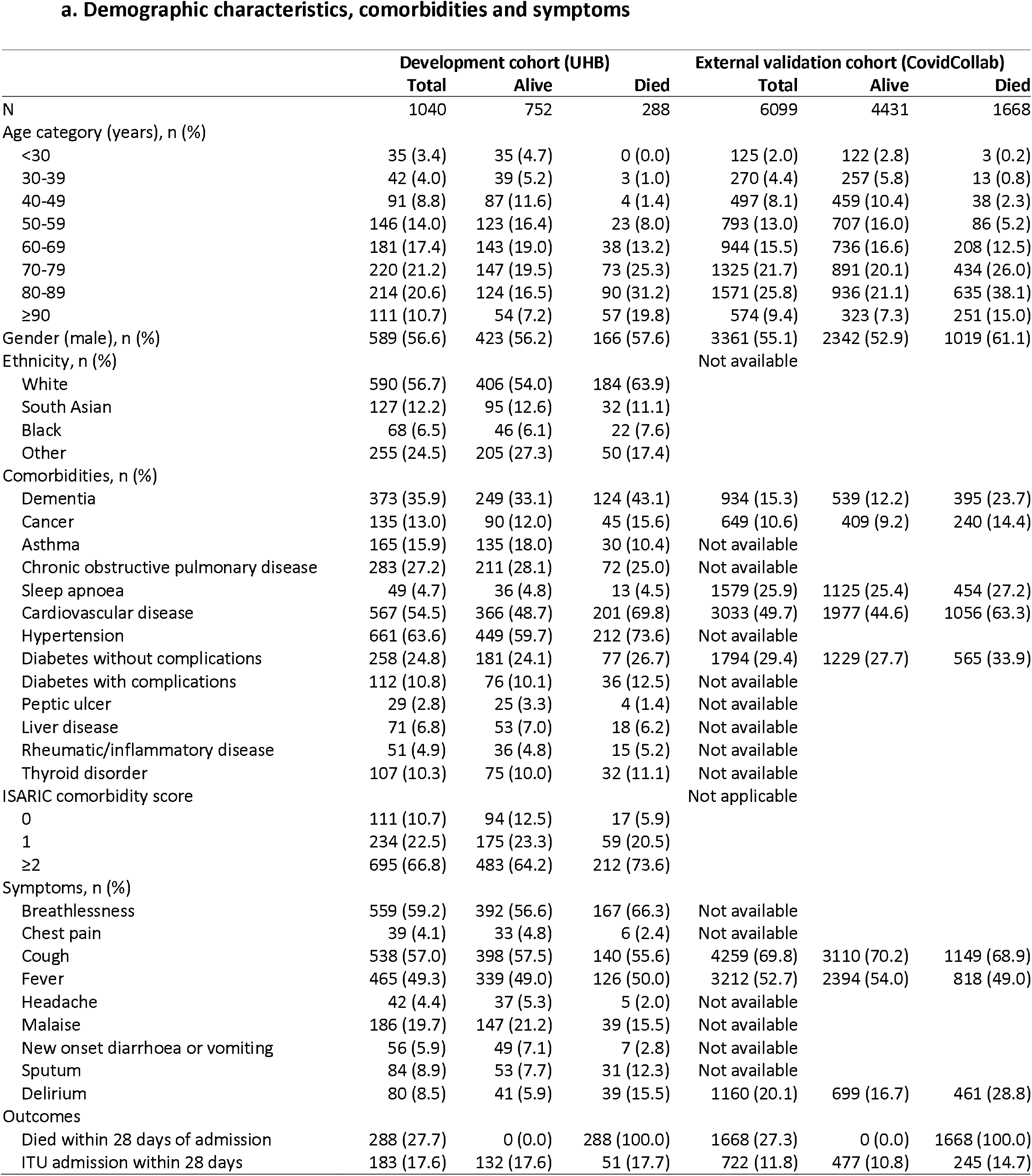
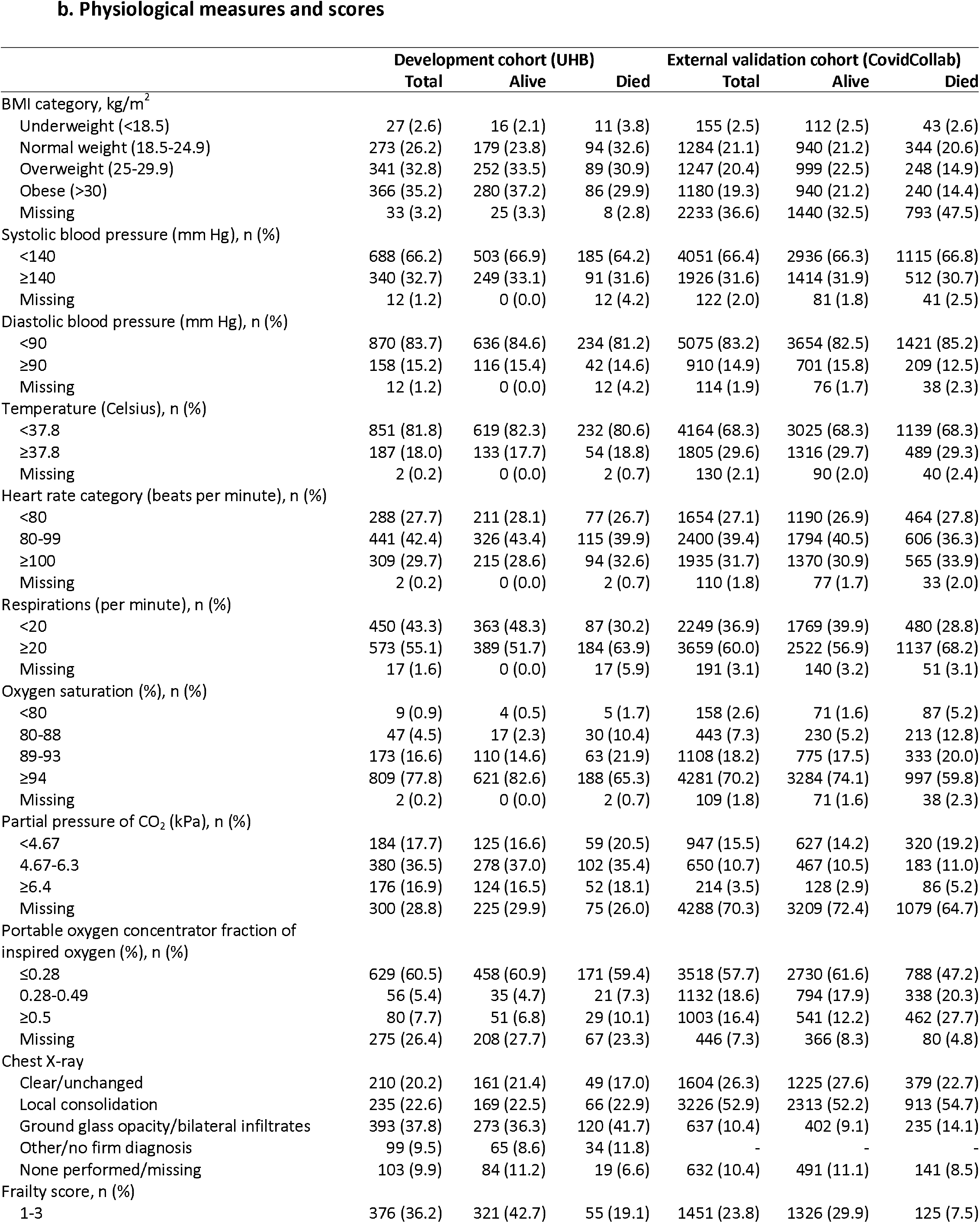
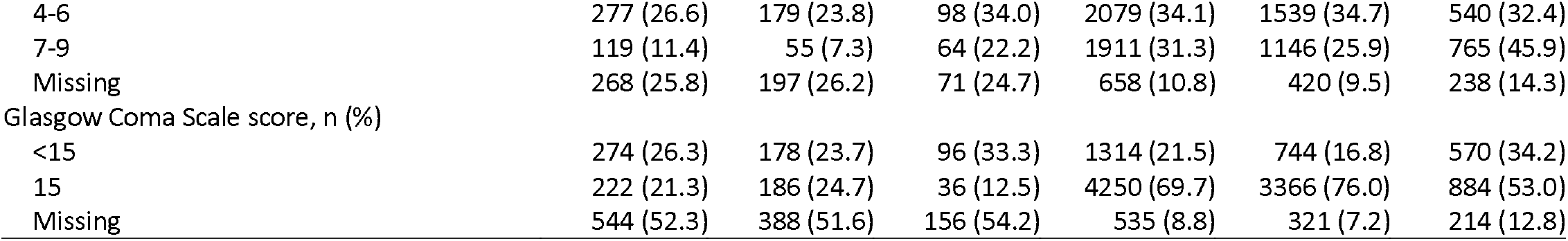
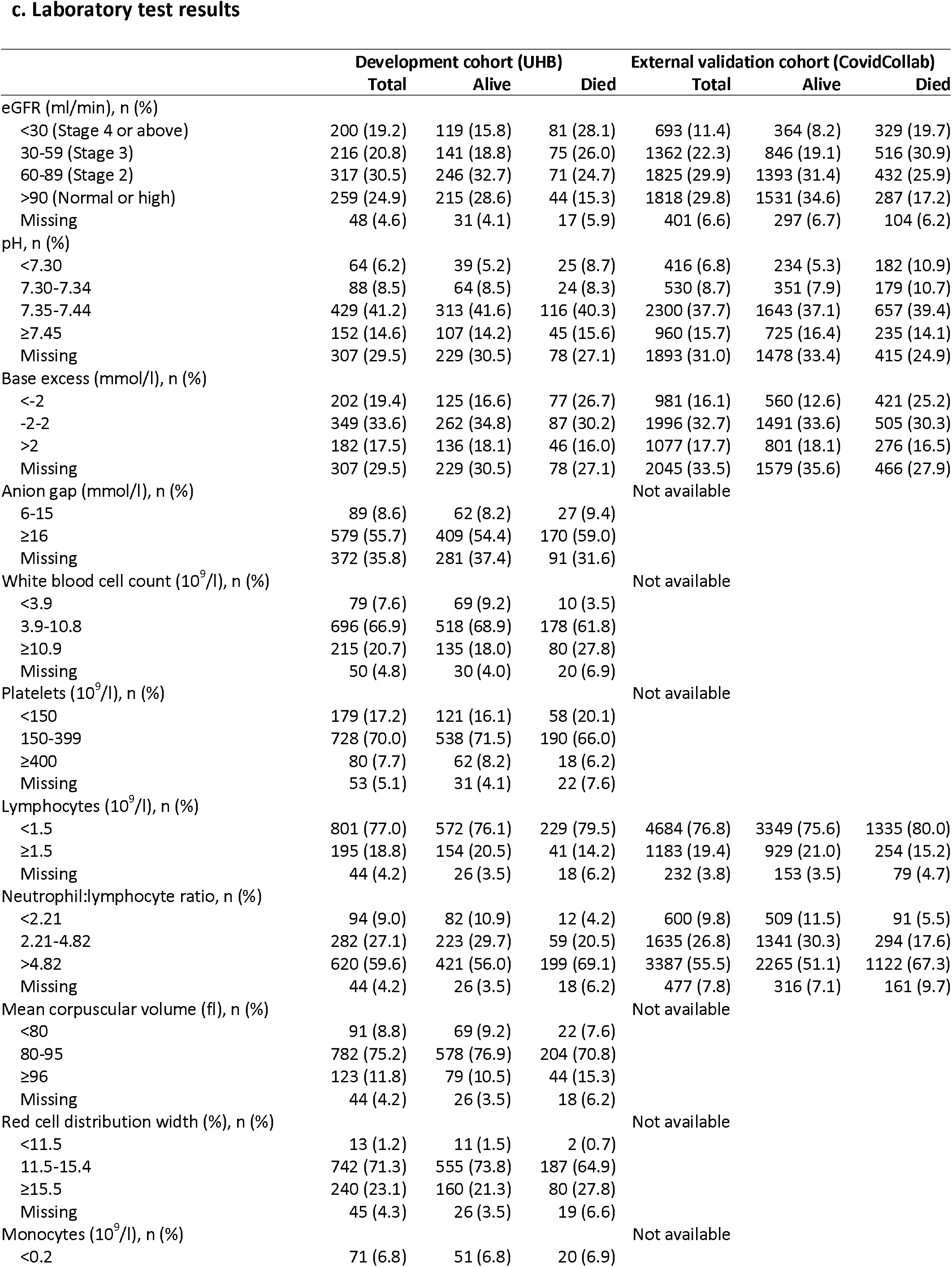
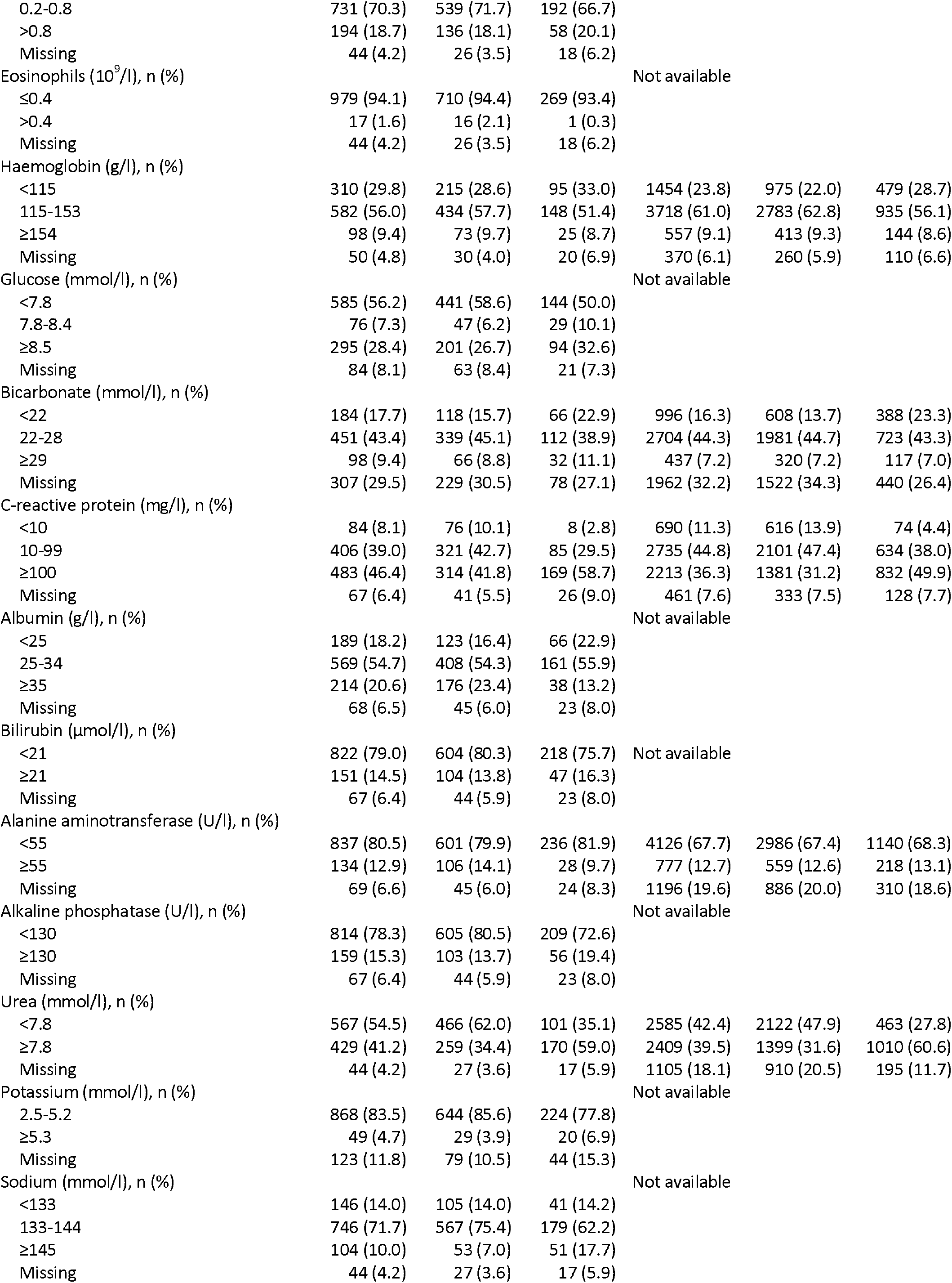
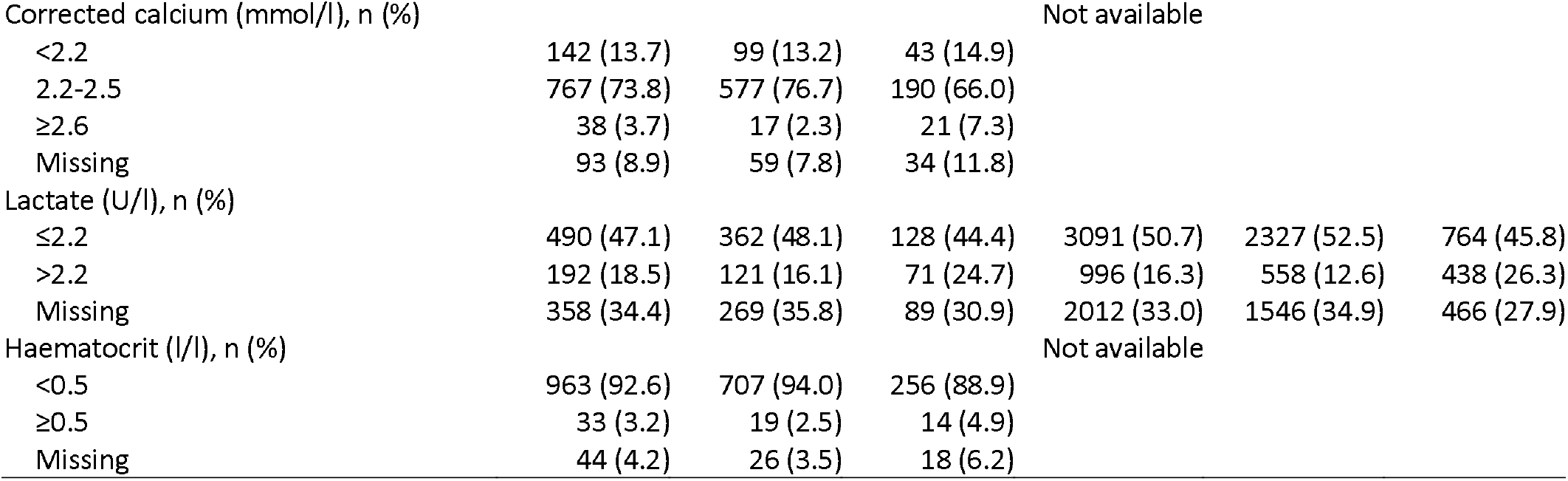
Baseline characteristics of participants admitted with COVID-19 in the derivation (University Hospitals Birmingham) and validation (CovidCollab) datasets.

### Candidate predictors

After exclusion of 7 candidate predictors with >40% missing data (D-dimer, ferritin, high sensitivity troponin, fibrinogen, lactate dehydrogenase, vitamin D, haemoglobin A1c), 63 predictors were considered for inclusion in the models:

Demographic characteristics: age, gender, ethnicity;

Symptoms (binary, presence or absence of symptom at admission): breathlessness, chest pain, cough, fever, headache, malaise, new onset diarrhoea or vomiting, sputum, delirium;

Physiological measures and vital signs: body mass index (BMI; kg/m^2^), systolic blood pressure (mm Hg), diastolic blood pressure (mm Hg), temperature (degrees Celsius), heart rate (beats per minute), respiratory rate (respirations per minute), oxygen saturation (%), partial pressure of CO_2_ (kPa), portable oxygen concentrator fraction of inspired oxygen (FiO_2_; %);

Imaging: chest X-ray finding (categorised as clear/unchanged, local consolidation, ground glass opacity/bilateral infiltrates, other/no firm diagnosis, none performed/missing);

Scores: Frailty score (Rockwood Clinical Frailty Scale);^12^ Glasgow Coma Scale score;^13,14^

Laboratory test results: estimated glomerular filtration rate (eGFR; ml/min), pH (%), base excess (mmol/l), anion gap (mmol/l), white blood cell (WBC) count (10^9^/l), platelets (10^9^/l), lymphocytes (10^9^/l), neutrophil:lymphocyte ratio, mean corpuscular volume (fl), red cell distribution width (%), monocytes (10^9^/l), eosinophils (10^9^/l), haemoglobin (g/l), glucose (mmol/l), bicarbonate (mmol/l), C-reactive protein (mg/l), albumin (g/l), bilirubin (μmol/l), alanine aminotransferase (U/l), alkaline phosphatase (U/l), urea (mmol/l), potassium (mmol/l), sodium (mmol/l), corrected calcium (mmol/l), lactate (U/l) and haematocrit (l/l);

Comorbidities (binary), presence or absence of record in discharge ICD-10 codes): dementia, cancer, asthma, chronic obstructive pulmonary disease, sleep apnoea, cardiovascular disease, hypertension, diabetes without complications, diabetes with complications, peptic ulcer, liver disease, rheumatic/inflammatory disease, thyroid disorder.

### 28-day mortality outcome: UHB model and predictive performance

Area under the ROC curve values for each of the logistic, LASSO and GBM models, treating continuous variables in one of three ways (as continuous variables with imputed missing values; as clinically meaningful categorical variables with imputed missing values; and as categorical variables with missing categories), are presented in Supplementary Table 1.

The final model selected was a logistic regression using stepwise selection of variables with categorisation of continuous variables (with imputed missing values). The final 18 categorical predictors included in the model were: age, breathlessness, sputum, systolic blood pressure, temperature, respiratory rate, oxygen saturation, FiO_2_, alkaline phosphatase, C-reactive protein, corrected calcium, eosinophils, glucose, pH, urea, WBC count, platelets, and frailty score. AUROC for the UHB cross-validated model was 0.778 (95 % CI 0.741-0.815) (Table 2). At a 20% predicted probability of mortality, sensitivity was 82% (95% CI 79-85), specificity was 59% (95% CI 56-61), positive predictive value was 43% (95% CI 41-45), and negative predictive was 90% (95% CI 88-91) (Table 3a). Calibration was very good at low to medium predicted probabilities, but was poorer at very high predicted probabilities; a calibration plot is shown in Figure 1A, calibration slope was 0.78 (95% CI 0.66-0.91) (Supplementary Table 2). Model coefficients (model equation) are shown in Supplementary Table 3.

**Table 2.** Area under the receiver operating characteristic curve (AUROC) for models developed in University Hospitals Birmingham (UHB) data and externally validated in CovidCollab data, and for external validation of the ISARIC model and 4C score.

| Dataset | Outcome | AUROC (95% CI) |
| --- | --- | --- |
| **Model development\*** |  |  |
| UHB | Mortality | 0.778 (0.741-0.815) |
| UHB | ITU admission | 0.892 (0.865-0.920) |
| UHB-R† | Mortality | 0.791 (0.761-0.822) |
| UHB-R† | ITU admission | 0.906 (0.883-0.929) |
| **External validation of new model\*** |  |  |
| CovidCollab | Mortality | 0.767 (0.754-0.780) |
| CovidCollab | ITU admission | 0.811 (0.795-0.828) |
| **External validation of ISARIC and 4C models in UHB data** |  |  |
| UHB – ISARIC model (XGBoost) | Mortality | 0.757 (0.721-0.792) |
| UHB – 4C score | Mortality | 0.754 (0.721-0.786) |

\*Models derived using logistic regression with stepwise selection of candidate predictors and categorisation of continuous variables into clinically meaningful categories (after imputing missing data).
†Not all variables included in the full University Hospital Birmingham (UHB) model were available in the CovidCollab dataset. Therefore, revised (reduced) models were developed in UHB data using a subset of the candidate predictors common to both the UHB and CovidCollab datasets (UHB-R), and these were then externally validated in the CovidCollab dataset.

**Table 3a.** Sensitivity, specificity, positive predictive value (PPV) and negative predictive value (NPV) for mortality at 28 days after admission (University Hospitals Birmingham derivation dataset)

| **Predicted probability** | **TP** | **TN** | **FP** | **FN** | **Sensitivity (95% CI)** | **Specificity (95% CI)** | **PPV (95% CI)** | **NPV (95% CI)** |
| --- | --- | --- | --- | --- | --- | --- | --- | --- |
| 10% | 271 | 277 | 475 | 17 | 0.941 (0.915-0.967) | 0.369 (0.355-0.382) | 0.363 (0.354-0.373) | 0.942 (0.917-0.968) |
| 20% | 236 | 441 | 311 | 52 | 0.821 (0.792-0.850) | 0.587 (0.563-0.611) | 0.432 (0.412-0.452) | 0.895 (0.878-0.912) |
| 30% | 194 | 544 | 208 | 94 | 0.673 (0.636-0.710) | 0.723 (0.703-0.743) | 0.482 (0.452-0.512) | 0.852 (0.836-0.869) |
| 40% | 152 | 626 | 126 | 136 | 0.527 (0.492-0.562) | 0.833 (0.815-0.850) | 0.547 (0.505-0.589) | 0.821 (0.807-0.835) |
| 50% | 118 | 680 | 72 | 170 | 0.410 (0.378-0.443) | 0.904 (0.888-0.919) | 0.620 (0.568-0.672) | 0.800 (0.789-0.811) |
| 60% | 82 | 713 | 39 | 206 | 0.284 (0.231-0.337) | 0.948 (0.944-0.952) | 0.675 (0.631-0.719) | 0.776 (0.763-0.788) |
| 70% | 50 | 728 | 24 | 238 | 0.174 (0.151-0.197) | 0.968 (0.967-0.969) | 0.673 (0.644-0.703) | 0.754 (0.748-0.759) |
| 80% | 24 | 741 | 11 | 264 | 0.082 (0.062-0.102) | 0.985 (0.978-0.993) | 0.685 (0.568-0.801) | 0.737 (0.733-0.741) |
| 90% | 8 | 750 | 2 | 280 | 0.027 (0.015-0.039) | 0.998 (0.995-1) | 0.808 (0.573-1) | 0.728 (0.725-0.731) |

TP = true positives, TN = true negatives, FP = false positives, FN = false negatives.

**Table 3b.**
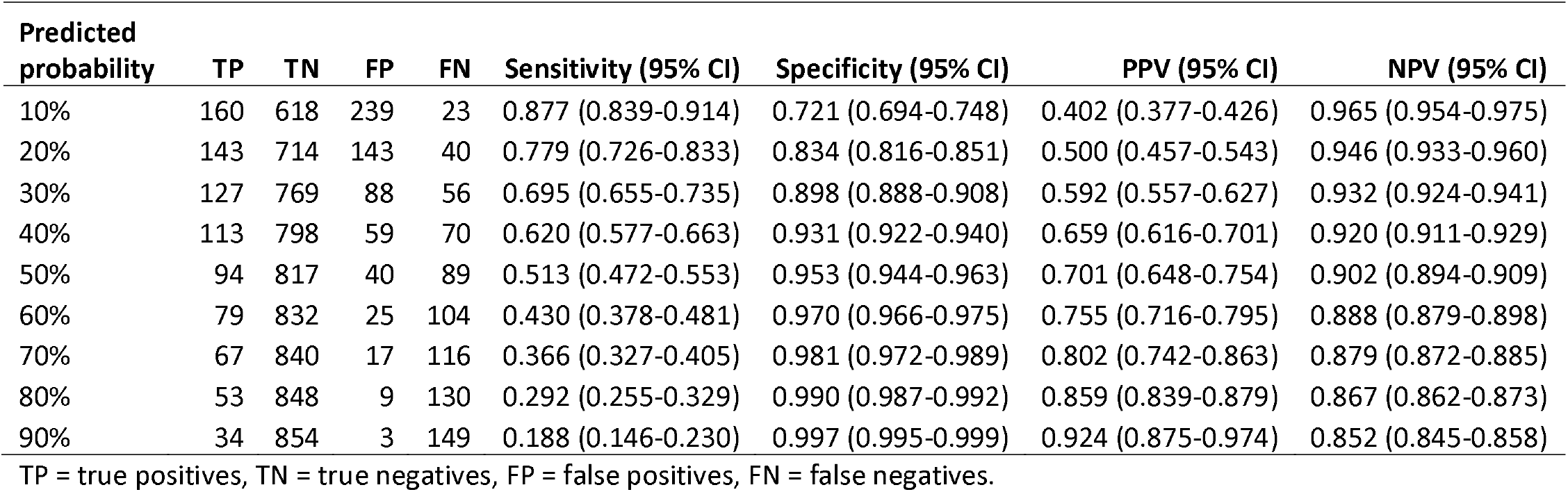
Sensitivity, specificity, positive predictive value (PPV) and negative predictive value (NPV) for intensive therapy unit admission within 28 days after admission (University Hospitals Birmingham derivation dataset)

| **Predicted probability** | **TP** | **TN** | **FP** | **FN** | **Sensitivity (95% CI)** | **Specificity (95% CI)** | **PPV (95% CI)** | **NPV (95% CI)** |
| --- | --- | --- | --- | --- | --- | --- | --- | --- |
| 10% | 160 | 618 | 239 | 23 | 0.877 (0.839-0.914) | 0.721 (0.694-0.748) | 0.402 (0.377-0.426) | 0.965 (0.954-0.975) |
| 20% | 143 | 714 | 143 | 40 | 0.779 (0.726-0.833) | 0.834 (0.816-0.851) | 0.500 (0.457-0.543) | 0.946 (0.933-0.960) |
| 30% | 127 | 769 | 88 | 56 | 0.695 (0.655-0.735) | 0.898 (0.888-0.908) | 0.592 (0.557-0.627) | 0.932 (0.924-0.941) |
| 40% | 113 | 798 | 59 | 70 | 0.620 (0.577-0.663) | 0.931 (0.922-0.940) | 0.659 (0.616-0.701) | 0.920 (0.911-0.929) |
| 50% | 94 | 817 | 40 | 89 | 0.513 (0.472-0.553) | 0.953 (0.944-0.963) | 0.701 (0.648-0.754) | 0.902 (0.894-0.909) |
| 60% | 79 | 832 | 25 | 104 | 0.430 (0.378-0.481) | 0.970 (0.966-0.975) | 0.755 (0.716-0.795) | 0.888 (0.879-0.898) |
| 70% | 67 | 840 | 17 | 116 | 0.366 (0.327-0.405) | 0.981 (0.972-0.989) | 0.802 (0.742-0.863) | 0.879 (0.872-0.885) |
| 80% | 53 | 848 | 9 | 130 | 0.292 (0.255-0.329) | 0.990 (0.987-0.992) | 0.859 (0.839-0.879) | 0.867 (0.862-0.873) |
| 90% | 34 | 854 | 3 | 149 | 0.188 (0.146-0.230) | 0.997 (0.995-0.999) | 0.924 (0.875-0.974) | 0.852 (0.845-0.858) |

TP = true positives, TN = true negatives, FP = false positives, FN = false negatives.

**Figure 1.**
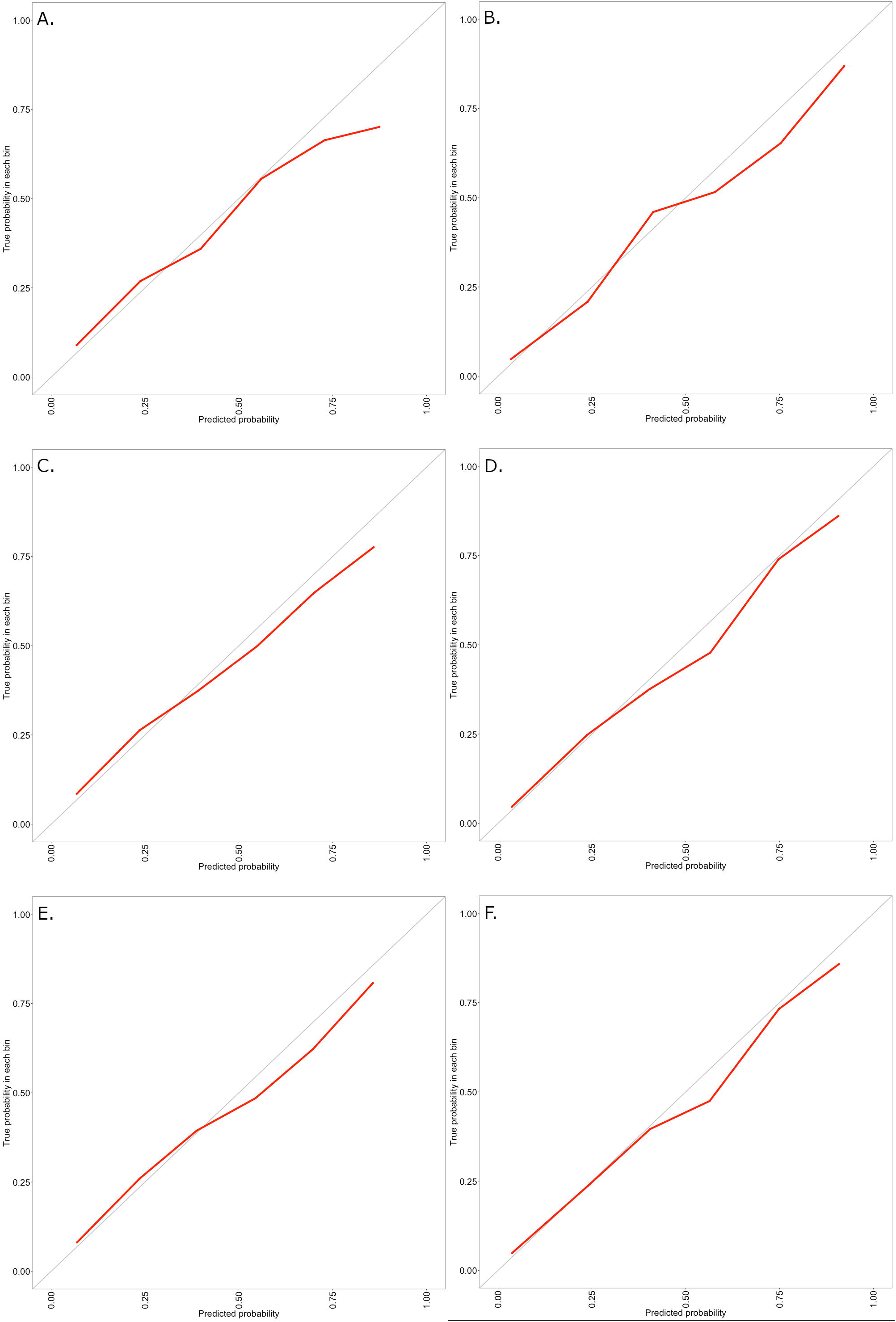
Calibration plots (observed probability (y-axis) against predicted probability (x-axis)): A. University Hospitals Birmingham (UHB) derivation dataset for mortality outcome; B. UHB derivation dataset for intensive therapy unit (ITU) admission outcome; C. UHB-R derivation/train dataset reduced model for mortality outcome; D. UHB-R derivation/train dataset reduced model for ITU admission outcome; E. CovidCollab external validation dataset reduced model for mortality outcome; F. CovidCollab external validation dataset reduced model for ITU admission outcome.

We used different methods of handling missing data in order to explore the impact on the model performance: in primary analysis, missing values were imputed; in secondary analysis, missing categories were used. The model derived using logistic stepwise selection using categorised variables with a missing category (rather than imputed data) gave a slightly higher AUROC (0.805, 95% CI 0.777-0.834); however, it was felt the inclusion of missing categories rather than imputed values might impact generalisability of the model outside the UHB/derivation data context. Other statistical modelling techniques (LASSO, GBM) offered no improvement in model performance and the models would be more difficult to implement in clinical practice. Using continuous predictors modelled using splines where non-linear relationship with the outcome was observed (Supplementary Figure 1) rather than categorising offered a small improvement in model performance (AUROC 0.791, 95% CI 0.759-0.823), but in the interests of clinical utility and interpretability the categorical model was favoured.

Addition of comorbidities to the candidate predictors included in the model did not improve performance of the model (Supplementary Table 1). Since comorbidities are known to be under-reported during acute presentations,^4^ and they offered no improvement on model performance, models without comorbidities were preferred.

### ITU admission: UHB model and predictive performance

Area under the ROC curve values for each of the models performed are presented in Supplementary Table 4.

**Table 4a.**
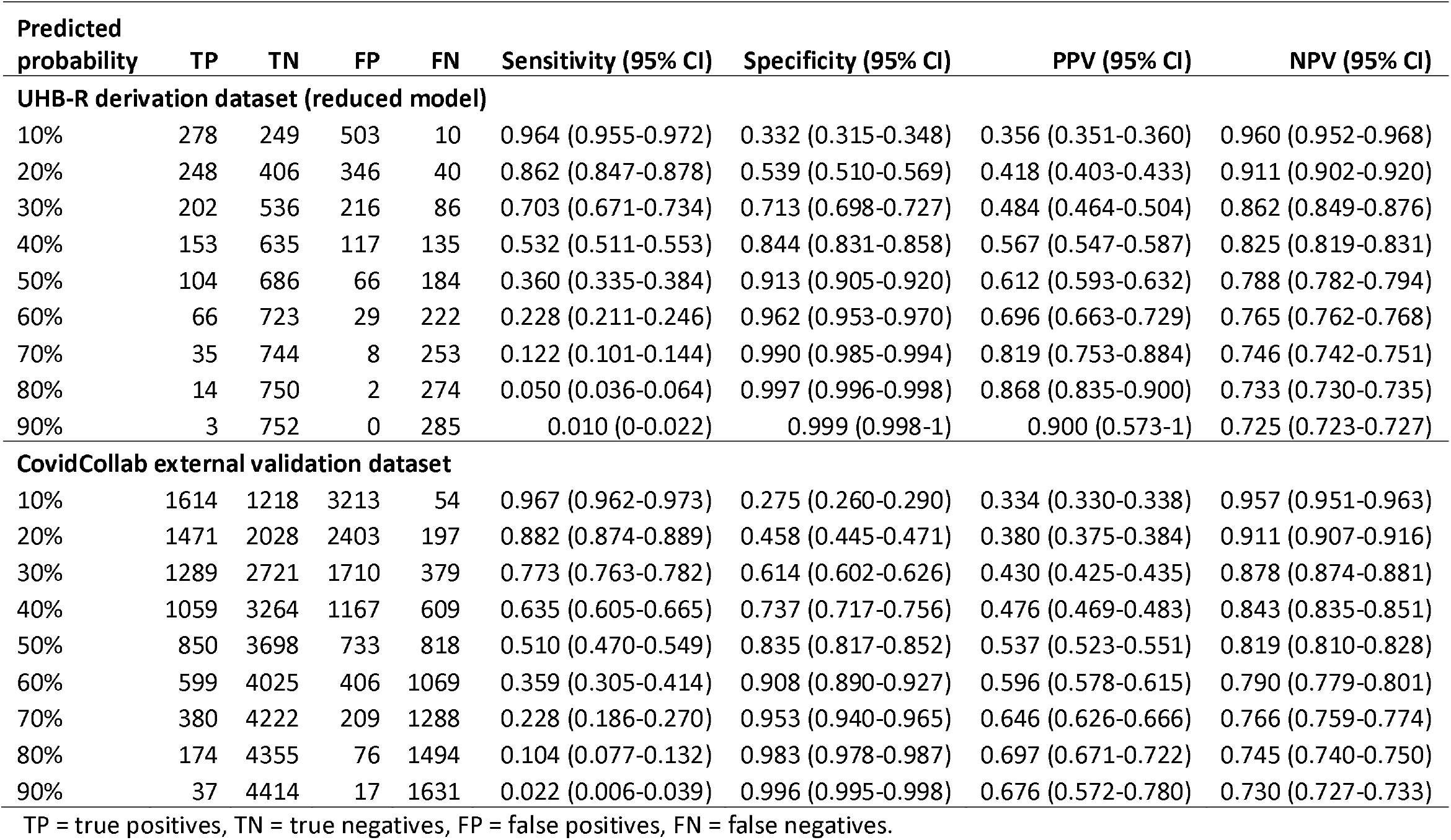
Sensitivity, specificity, positive predictive value (PPV) and negative predictive value (NPV) for mortality at 28 days after admission (University Hospitals Birmingham derivation dataset and CovidCollab external validation dataset, using predictors common to both datasets)

| **Predicted probability** | **TP** | **TN** | **FP** | **FN** | **Sensitivity (95% CI)** | **Specificity (95% CI)** | **PPV (95% CI)** | **NPV (95% CI)** |
| --- | --- | --- | --- | --- | --- | --- | --- | --- |
| **UHB-R derivation dataset (reduced model)** |  |  |  |  |  |  |  |  |
| 10% | 278 | 249 | 503 | 10 | 0.964 (0.955-0.972) | 0.332 (0.315-0.348) | 0.356 (0.351-0.360) | 0.960 (0.952-0.968) |
| 20% | 248 | 406 | 346 | 40 | 0.862 (0.847-0.878) | 0.539 (0.510-0.569) | 0.418 (0.403-0.433) | 0.911 (0.902-0.920) |
| 30% | 202 | 536 | 216 | 86 | 0.703 (0.671-0.734) | 0.713 (0.698-0.727) | 0.484 (0.464-0.504) | 0.862 (0.849-0.876) |
| 40% | 153 | 635 | 117 | 135 | 0.532 (0.511-0.553) | 0.844 (0.831-0.858) | 0.567 (0.547-0.587) | 0.825 (0.819-0.831) |
| 50% | 104 | 686 | 66 | 184 | 0.360 (0.335-0.384) | 0.913 (0.905-0.920) | 0.612 (0.593-0.632) | 0.788 (0.782-0.794) |
| 60% | 66 | 723 | 29 | 222 | 0.228 (0.211-0.246) | 0.962 (0.953-0.970) | 0.696 (0.663-0.729) | 0.765 (0.762-0.768) |
| 70% | 35 | 744 | 8 | 253 | 0.122 (0.101-0.144) | 0.990 (0.985-0.994) | 0.819 (0.753-0.884) | 0.746 (0.742-0.751) |
| 80% | 14 | 750 | 2 | 274 | 0.050 (0.036-0.064) | 0.997 (0.996-0.998) | 0.868 (0.835-0.900) | 0.733 (0.730-0.735) |
| 90% | 3 | 752 | 0 | 285 | 0.010 (0-0.022) | 0.999 (0.998-1) | 0.900 (0.573-1) | 0.725 (0.723-0.727) |
| **CovidCollab external validation dataset** |  |  |  |  |  |  |  |  |
| 10% | 1614 | 1218 | 3213 | 54 | 0.967 (0.962-0.973) | 0.275 (0.260-0.290) | 0.334 (0.330-0.338) | 0.957 (0.951-0.963) |
| 20% | 1471 | 2028 | 2403 | 197 | 0.882 (0.874-0.889) | 0.458 (0.445-0.471) | 0.380 (0.375-0.384) | 0.911 (0.907-0.916) |
| 30% | 1289 | 2721 | 1710 | 379 | 0.773 (0.763-0.782) | 0.614 (0.602-0.626) | 0.430 (0.425-0.435) | 0.878 (0.874-0.881) |
| 40% | 1059 | 3264 | 1167 | 609 | 0.635 (0.605-0.665) | 0.737 (0.717-0.756) | 0.476 (0.469-0.483) | 0.843 (0.835-0.851) |
| 50% | 850 | 3698 | 733 | 818 | 0.510 (0.470-0.549) | 0.835 (0.817-0.852) | 0.537 (0.523-0.551) | 0.819 (0.810-0.828) |
| 60% | 599 | 4025 | 406 | 1069 | 0.359 (0.305-0.414) | 0.908 (0.890-0.927) | 0.596 (0.578-0.615) | 0.790 (0.779-0.801) |
| 70% | 380 | 4222 | 209 | 1288 | 0.228 (0.186-0.270) | 0.953 (0.940-0.965) | 0.646 (0.626-0.666) | 0.766 (0.759-0.774) |
| 80% | 174 | 4355 | 76 | 1494 | 0.104 (0.077-0.132) | 0.983 (0.978-0.987) | 0.697 (0.671-0.722) | 0.745 (0.740-0.750) |
| 90% | 37 | 4414 | 17 | 1631 | 0.022 (0.006-0.039) | 0.996 (0.995-0.998) | 0.676 (0.572-0.780) | 0.730 (0.727-0.733) |

TP = true positives, TN = true negatives, FP = false positives, FN = false negatives.

**Table 4b.** Sensitivity, specificity, positive predictive value (PPV) and negative predictive value (NPV) for intensive therapy unit admission within 28 days after admission (University Hospitals Birmingham derivation dataset and CovidCollab external validation dataset, using predictors common to both datasets)

| Predicted probability | TP | TN | FP | FN | Sensitivity (95% CI) | Specificity (95% CI) | PPV (95% CI) | NPV (95% CI) |
| --- | --- | --- | --- | --- | --- | --- | --- | --- |
| **UHB-R derivation dataset (reduced model)** |  |  |  |  |  |  |  |  |
| 10% | 165 | 590 | 267 | 18 | 0.904 (0.877-0.931) | 0.689 (0.671-0.707) | 0.383 (0.368-0.398) | 0.971 (0.963-0.979) |
| 20% | 152 | 709 | 148 | 31 | 0.831 (0.812-0.849) | 0.827 (0.816-0.839) | 0.507 (0.494-0.519) | 0.958 (0.954-0.962) |
| 30% | 132 | 765 | 92 | 51 | 0.723 (0.698-0.749) | 0.893 (0.882-0.904) | 0.591 (0.567-0.615) | 0.938 (0.933-0.943) |
| 40% | 112 | 803 | 54 | 71 | 0.613 (0.573-0.653) | 0.937 (0.928-0.946) | 0.675 (0.649-0.702) | 0.919 (0.912-0.926) |
| 50% | 92 | 829 | 28 | 91 | 0.505 (0.483-0.526) | 0.967 (0.956-0.979) | 0.768 (0.708-0.829) | 0.901 (0.898-0.905) |
| 60% | 77 | 841 | 16 | 106 | 0.419 (0.403-0.435) | 0.981 (0.974-0.988) | 0.828 (0.779-0.877) | 0.888 (0.886-0.890) |
| 70% | 67 | 847 | 10 | 116 | 0.365 (0.344-0.386) | 0.988 (0.983-0.994) | 0.870 (0.816-0.924) | 0.879 (0.876-0.883) |
| 80% | 46 | 851 | 6 | 137 | 0.251 (0.189-0.313) | 0.993 (0.989-0.997) | 0.889 (0.840-0.938) | 0.861 (0.852-0.871) |
| 90% | 20 | 855 | 2 | 163 | 0.108 (0.067-0.149) | 0.997 (0.996-0.999) | 0.898 (0.838-0.957) | 0.840 (0.833-0.846) |
| **CovidCollab external validation dataset** |  |  |  |  |  |  |  |  |
| 10% | 574 | 3623 | 1754 | 148 | 0.794 (0.764-0.825) | 0.674 (0.651-0.697) | 0.247 (0.238-0.255) | 0.961 (0.956-0.965) |
| 20% | 465 | 4312 | 1065 | 257 | 0.644 (0.621-0.667) | 0.802 (0.785-0.819) | 0.304 (0.288-0.321) | 0.944 (0.941-0.947) |
| 30% | 377 | 4655 | 722 | 345 | 0.523 (0.497-0.549) | 0.866 (0.847-0.885) | 0.344 (0.320-0.367) | 0.931 (0.929-0.933) |
| 40% | 315 | 4881 | 496 | 407 | 0.437 (0.381-0.492) | 0.908 (0.887-0.928) | 0.389 (0.361-0.417) | 0.923 (0.918-0.929) |
| 50% | 250 | 5045 | 332 | 472 | 0.346 (0.258-0.434) | 0.938 (0.920-0.956) | 0.430 (0.413-0.447) | 0.914 (0.905-0.923) |
| 60% | 178 | 5168 | 209 | 544 | 0.247 (0.175-0.318) | 0.961 (0.945-0.977) | 0.462 (0.420-0.503) | 0.905 (0.898-0.912) |
| 70% | 113 | 5256 | 121 | 609 | 0.157 (0.105-0.208) | 0.978 (0.967-0.988) | 0.486 (0.439-0.534) | 0.896 (0.891-0.901) |
| 80% | 57 | 5319 | 58 | 665 | 0.079 (0.044-0.114) | 0.989 (0.984-0.995) | 0.500 (0.410-0.590) | 0.889 (0.886-0.892) |
| 90% | 18 | 5357 | 20 | 704 | 0.025 (0.006-0.044) | 0.996 (0.993-1) | 0.485 (0.367-0.603) | 0.884 (0.882-0.886) |

TP = true positives, TN = true negatives, FP = false positives, FN = false negatives.

The final model selected was a logistic regression using stepwise selection of variables with categorisation of continuous variables (with imputed missing values). The final 16 categorical predictors included in the model were: age, gender, fever, new onset diarrhoea or vomiting, heart rate, respiratory rate, FiO_2_, temperature, albumin, C-reactive protein, eGFR, pH, monocytes, WBC, frailty score, and Glasgow Coma Scale score. AUROC was 0.892 (95% CI 0.865-0.920) (Table 2). At a 20% predicted probability of ITU admission, sensitivity was 78% (95% CI 73-83), specificity was 83% (95% CI 82-85), positive predictive value was 50% (95% CI 46-54), and negative predictive was 95% (95% CI 93-96) (Table 3b). Calibration was good; a calibration plot is shown in Figure 1B, calibration slope was 0.89 (95% CI 0.77-1.02) (Supplementary Table 2). Model coefficients are shown in Supplementary Table 5.

Again, use of a missing category rather than imputed missing values improved model performance but was not selected as the final model due to potential problems with generalisability (AUROC 0.955, 95% CI 0.933-0.978). Models using continuous rather than categorical predictors (with splines where relevant; Supplementary Figure 2) gave a small improvement in AUROC (0.908, 95% CI 0.883-0.934), but were deemed less practical. Addition of comorbidities to the predictors included in the model did not improve performance.

### Reduced UHB model and external validation in the CovidCollab dataset

A total of 6099 patients admitted with COVID-19 were included in the CovidCollab external validation dataset; 1668 (27%) died and 722 (12%) were admitted to ITU. Not all variables included in the UHB model derived above were available in the CovidCollab dataset. Therefore, revised and reduced models were developed in UHB data using the subset of candidate predictors common to both the UHB and CovidCollab datasets (reduced UHB dataset, UHB-R), using logistic regression with stepwise selection, and these were then externally validated in the CovidCollab dataset. The reduced set of 27 candidate predictors included: demographic characteristics: age, gender; symptoms: cough, fever, delirium; physiological measures and vital signs: BMI, systolic blood pressure, diastolic blood pressure, heart rate, temperature, respiratory rate, oxygen saturation, FiO_2_, chest X-ray; frailty score; Glasgow Coma Scale score; laboratory test results: eGFR, pH, base excess, lymphocytes, neutrophil:lymphocyte ratio, haemoglobin, bicarbonate, C-reactive protein, alanine aminotransferase, urea, and lactate.

For the 28-day mortality outcome, following stepwise selection, the final 10 categorical predictors (common to both datasets) included in the logistic regression model were: age, oxygen saturation, FiO_2_, respiratory rate, temperature, systolic blood pressure, C-reactive protein, pH, urea, and frailty score. These predictors were a subset of those in the original UHB model derivation, but gave similar model performance. AUROC in the UHB-R dataset was 0.791 (95% CI 0.761-0.822), and in the CovidCollab dataset was 0.767 (95% CI 0.754-0.780) (Table 2). At a 20% predicted probability of mortality, in the UHB-R dataset sensitivity was 86% (95% CI 85-88), specificity was 54% (95% CI 51-57), PPV was 42% (95% CI 40-43) and NPV was 91% (95% CI 90-92); in the CovidCollab dataset sensitivity was 88% (95% CI 87-89), specificity was 46% (95% CI 45-47), PPV was 38% (95% CI 0.37-0.38), and NPV was 91% (95% CI 91-92) (Table 4a). Calibration was good for both derivation and external validation datasets; calibration plots are shown in Figures 1C-D, calibration slopes were 0.86 (95% CI 0.82-0.90) and 0.88 (95% CI 0.80-0.96) for the UHB-R and CovidCollab datasets, respectively (Supplementary Table 2). Model coefficients are shown in Supplementary Table 6.

For the ITU admission outcome, the final 11 categorical predictors (common to both datasets) included in the model were: age, gender, fever, respiratory rate, FiO_2_, C-reactive protein, eGFR, pH, neutrophil:lymphocyte ratio, frailty score, and Glasgow Coma Scale score. AUROC in the UHB-R dataset was 0.906 (95% CI 0.883-0.929), and in the CovidCollab dataset was 0.811 (95% CI 0.795-0.828) (Table 2). At a 20% predicted probability of ITU admission, in the UHB-R dataset sensitivity was 83% (95% CI 81-85), specificity was 83% (95% CI 82-84), PPV was 51% (95% CI 49-52) and NPV was 96% (95% CI 95-96); in the CovidCollab dataset sensitivity was 64% (95% CI 62-67), specificity was 80% (95% CI 79-82), PPV was 30% (95% CI29-32), and NPV was 94% (95% CI 94-95) (Table 4b). Calibration was good for both derivation and external validation datasets; calibration plots are shown in Figures 1E-F, calibration slopes were 0.94 (95% CI 0.84-1.03) and 0.93 (95% CI 0.84-1.02) for the UHB-R and CovidCollab datasets, respectively (Supplementary Table 2). Model coefficients are shown in Supplementary Table 7.

### External validation of the ISARIC models in the UHB dataset

The recently published ISARIC model (XGBoost) was recalibrated to the UHB data and performance in the UHB data assessed for predicting mortality; AUROC was 0.757 (95% CI 0.721-0.792) (Table 2). For the simplified 4C score (logistic regression), the AUROC was marginally lower at 0.754 (95% CI 0.721-0.786).

It was not possible to externally validate the ISARIC model and 4C score in the CovidCollab dataset, as information on many of the comorbidities required to calculate the ISARIC comorbidity score was not available in the dataset.

### Sensitivity analyses

Sensitivity analyses were carried out using the logistic regression models initially derived in the UHB dataset (18 predictors for mortality outcome and 16 predictors for ITU admission).

### Complete case analysis

Few patients in the dataset had complete data (n = 224/1040, 22%); model performance in this patient subset was slightly poorer for mortality outcome: AUROC 0.690 (95% CI 0.588-0.791) for mortality and 0.887 (95% CI 0.825-0.949) for ITU admission. Including patients with missing variables, with missing values imputed, improved model performance; allowing even a single missing/imputed variable improved AUROC for mortality to 0.767 (95% CI 0.715-0.819) (Supplementary Table 8).

### Stratification by gender

When patients were stratified by gender, the model predicting mortality performed slightly worse in males (AUROC 0.766, 95% CI 0.700-0.831) than females (0.788, 95% CI 0.729-0.847). The model predicting ITU admission performed similarly in males and females: 0.893 (95% CI 0.823-0.963) in males and 0.893 (95% CI 0.851-0.935) in females (Supplementary Table 8). Training models in gender-specific datasets offered no improvement on model performance (Supplementary Table 9).

### Time series analysis

Supplementary Figure 3 shows variation in logistic regression coefficients for the candidate predictors from day of admission and up to 7 days later. The majority of coefficients remained relatively constant over time. However, several (not necessarily statistically significant) trends in the modification of effects over the week of admission on mortality were visible, such as a decrease over the week of the effect of obesity on mortality, elevated effect of eosinophils, and an increase over the week of the effect of elevated haemoglobin, elevated potassium and elevated oxygen saturation. Some of these might be depletion effects related to relatively high patient mortality in the first few days, for example, the apparent protective effect of obesity and high eosinophils.

## Discussion

Using routinely collected data for more than a thousand patients admitted with COVID-19 at a large UK hospital trust, we have developed and externally validated prognostic models for mortality and ITU admission. The models showed good discrimination and calibration. The candidate predictors explored included a clinically informed, wider range of demographics, clinical observations, symptoms, comorbidities, biomarkers and radiological investigations than those included in the derivation of existing prognostic scores or models. If integrated into hospital electronic medical records systems, the model algorithms will provide a predicted probability of mortality or ITU admission within 28 days of hospital admission for each patient based on their individual data at, or close to, the time of admission, which will support clinicians’ decision making with regard to appropriate patient care pathways and triage. This information might also assist clinicians in explaining complex prognostic assessments and decisions to patients and their relatives, particularly at a time when relatives are unable to see the patient and understand how unwell they are.

The models developed using all 63 available candidate predictors from UHB performed well with an optimism-adjusted AUROC of 0.778 (95 % CI 0.741-0.815) for mortality within 28 days of admission and 0.892 (95% CI 0.865-0.920) for ITU admission. Models performed well in male and female subgroups.

Not all variables included in the UHB dataset are routinely collected at admission in other hospitals; therefore, reduced models using only variables common to both UHB and the CovidCollab external validation dataset were explored. These were found to have slightly better discrimination, with an AUROC of 0.791 (95% CI 0.761-0.822) for mortality and 0.906 (95% CI 0.883-0.929) for ITU admission in the UHB derivation dataset. These reduced models also performed well in the CovidCollab external validation dataset, with AUROCs of 0.767 (95% CI 0.754-0.780) and 0.811 (95% CI 0.795-0.828) for mortality and ITU admission, respectively. Calibration of all models showed good agreement between observed and predicted probabilities, particularly at lower predicted probabilities in the range where the models would be of most clinical utility.

Two systematic reviews summarised the existing secondary care COVID-19 prognostic models or scores published until the 31^st^ May 2020.^1,15^ Many of the reported models (largely derived from Chinese cohorts) demonstrated high discriminatory performance, however all pre-existing models when assessed using the PROBAST score were at high risk of bias. Additionally, few models were externally validated in suitable cohorts. By deriving our model from routinely collected data, we were able to reduce the risk of bias in patient selection as well as predictor and outcome measurements. Additionally, in this study we were able to externally validate models in a large global heterogeneous cohort. Our study builds on the existing literature as the CovidCollab dataset includes some patients from low-middle income countries where validation studies of prediction models are yet to be published.

More recently, the most notable secondary care prediction model advised for uptake in UK hospitals was derived from the ISARIC-WHO collaborating cohort.^3^ Both the full and reduced UHB-derived models for mortality had slightly better discrimination than the ISARIC model and 4C score in the UHB data (AUROC 0.757 (95% CI 0.724-0.791) for recalibrated ISARIC model and 0.754 (95% CI 0.721-0.786) for the simplified 4C score).This compares with AUROCs of 0.779 (0.772 to 0.785) for the ISARIC model and 0.767 (0.760 to 0.773) for the 4C score reported in the original ISARIC validation cohort.^3^ Furthermore, the newly developed UHB model offers an advantage over the ISARIC and 4C models in that it utilises only routinely collected patient data recorded at admission, and does not require additional assessment and recording of specific comorbidities.

In our time series analysis we did not find strong evidence for trends in predictor coefficients over the first 8 days of admission, particularly for variables included in the final models, suggesting that time-dependent effects due to effect modification or selection bias in the first week are small. If models were implemented to treat time after admission in a fine-grained way, only a limited subset of biomarkers might be of interest. Another recent model derived from patients with COVID-19 in a Hong Kong hospital adopted the use of time-dependent routinely collected predictors; the model in the Hong Kong study demonstrated high discrimination, with an AUROC of 0.91 when predicting severe COVID-19 outcomes.^16^ However, this model is yet to be externally validated.

### Strengths and limitations

The UHB dataset represents one of the largest and most ethnically diverse patient cohorts within the UK. Additionally, as part of the early UHB response to the COVID-19 pandemic, the hospital trust ensured that, upon admission, all patients underwent a wide range of investigations to support international research efforts examining prognostic markers. This allowed us to examine a wide range of possible predictors (63 candidate predictors after exclusions). Lastly, a strength of this study was the good performance, in terms of both discrimination and calibration, of the simplified, reduced model in an externally validated cohort (CovidCollab), indicating its suitability for wider use, including potentially in LMICs.

Despite the strengths, the findings must be considered in light of the study’s limitations. Although we were able to use a derivation dataset from UHB with low levels of missing data, the overall sample size was relatively small compared to that of the ISARIC study and was limited to one UK geographical location. However, we were able to externally validate the model in a larger external cohort. A second limitation was that in the external validation cohort we were unable to examine all of the predictors included in the original full UHB model due to only a reduced set of candidate predictors being available in CovidCollab. Nevertheless, the model performed well and the results suggest it may be applicable in a wide range of datasets where only a reduced set of predictor variables is available. It was not possible to carry out stratified analysis by ethnicity as, in the UHB dataset, too few patients were included in most of the strata; ethnicity data was not available in the CovidCollab dataset.

## Conclusion

In this paper we have described the development and external validation of novel prognostic models which predict mortality and ITU admission within 28 days of admission for patients admitted to hospital with COVID-19, using routinely collected data gathered at admission. The simple, reduced models showed good discrimination and calibration, outperformed the existing ISARIC model and 4C score, and performed well in a validation cohort. The models can be integrated into existing electronic medical records systems to calculate each individual patient’s probability of death or ITU admission at the time of hospital admission. The models should be further validated to determine their applicability in other populations. In addition, implementation of the models and clinical utility should be evaluated.

## Supporting information

Tripod checklist

## Data Availability

No additional data available.

## Ethical approval

Ethical approval was provided by the East Midlands – Derby REC (reference: 20/EM/0158) for the PIONEER Research Database (data from University Hospitals Birmingham).

For CovidCollab data, local, regional, and national approvals were obtained from all participating sites. In the UK, this study was registered as clinical audit or service evaluation, with approval granted in line with local information governance policies, in line with assessment and guidance by the Health Research Authority. At the lead site (University Hospitals Birmingham NHS Trust) this study was registered as clinical audit (CARMS-15986). In other countries, local principal investigators were responsible for obtaining approvals in line with their local, regional, and national guidelines and recommendations. Only routinely collected data was collected and patient care was not altered by this study. Anonymised data was securely transferred to the Birmingham Centre for Prospective and Observational Studies (BiCOPS), University of Birmingham via REDCap. All sites were required to confirm that approvals were in place prior to being provided with logins; written data sharing agreements were arranged where requested by individual sites.

### Contributorship statement

The study was designed by NA, KN, ES, MP, YT, CS and TT. Data was collected by KG, ES, CS and SG. TT, NA and DG analysed the data. The first draft was written by NA and TT. All authors critically reviewed and revised the manuscript.

## Transparency declaration

The lead author (the manuscript’s guarantor) affirms that this manuscript is an honest, accurate, and transparent account of the study being reported; that no important aspects of the study have been omitted; and that any discrepancies from the study as planned (and, if relevant, registered) have been explained.

## Exclusive licence

The Corresponding Author has the right to grant on behalf of all authors and does grant on behalf of all authors, a worldwide licence (http://www.bmj.com/sites/default/files/BMJ%20Author%20Licence%20March%202013.doc) to the Publishers and its licensees in perpetuity, in all forms, formats and media (whether known now or created in the future), to i) publish, reproduce, distribute, display and store the Contribution, ii) translate the Contribution into other languages, create adaptations, reprints, include within collections and create summaries, extracts and/or, abstracts of the Contribution and convert or allow conversion into any format including without limitation audio, iii) create any other derivative work(s) based in whole or part on the on the Contribution, iv) to exploit all subsidiary rights to exploit all subsidiary rights that currently exist or as may exist in the future in the Contribution, v) the inclusion of electronic links from the Contribution to third party material where-ever it may be located; and, vi) licence any third party to do any or all of the above.

## Data sharing

No additional data available.

## Dissemination declaration

Dissemination to participants is not possible as this is a review of existing evidence. Patients and the public will not be involved in dissemination of the findings.

## Acknowledgements

MP and YT are supported by the NIHR Birmingham Biomedical Research Centre. The views expressed are those of the author(s) and not necessarily those of the NHS, NIHR or the Department of Health and Social Care.

We would also like to acknowledge the collaborators for the CovidCollab study: Sarah Richardson, Miles Witham, AGE Research Group, NIHR Newcastle Biomedical Research Centre, University of Newcastle and Newcastle-upon-Tyne Hospitals NHS Foundation Trust, UK; Omar A Abdelwahab, Elsayed M Awad, Ahmed Y Azzam, Ahmed Cordie, Ahmed O Elmehrath, Mostafa El-Shazly, Almontacer EB Masood, Alazher University Hospital, Egypt; Osama MAS Abdulhadi, Hazem Ahmed, Muhammed Elhadi, Ahmed KM Hadreiez, Ahmed A Momen, Mosab AA Shaban, Alkhadra Hospital, Libya; Giuseppe Cecere, Aldo Rocca, Antonio Cardarelli Hospital, Italy; Hossam Aldein S Abd Elazeem, Mohammed H Abdelhafez, Islam A Ahmed, Shrouk M Elghazaly, Helal F Hetta, Mohamed Eltaher AA Ibrahim, Soha M Mohamed, Aliae AR Mohamed Hussein, Mohamed M Moustafa, Mariam Albatoul Nageh, Mahmoud M Saad, Alshaimaa M Saad, Omar Zein Elabedeen, Assiut University Hospital, Egypt; Victoria Cox, Danielle Hunsley, Rebecca Ryall, Kathleen T Shakespeare, Thyn Thyn, Rachael Webb, Barnsley Hospital NHS Foundation Trust, UK; Deepthy Hari Madhavan, Nik Sanyal, Birmingham Heartlands Hospital, UK; Bryony Brown, Matthew Hale, Bradford Teaching Hospitals NHS Foundation Trust, UK; Marie Goujon, Cambridge University Hospitals NHS Foundation Trust, UK; Benjamin Jelley, Cardiff University, UK; Laxmi Babar, Tina Doll, Agnieszka Felska, Daniel N Guerero, Sandeep Karthikeyan, Anne Karunatilleke, Helena Lee, Emma Livesey, Amelia Roberts, Charlotte Roberts-Rhodes, City Hospital, Birmingham, UK; Teresa Perra, Alberto Porcu, Cliniche San Pietro, A.O.U. Sassari, Italy; Antonio Buondonno, Giuseppe Cecere, Aldo Rocca, Department of Medicine and Health Science “V. Tiberio”, University of Molise, Italy; Vesna Hogan, Iain Wilkinson, East Surrey Hospital, UK; Ioannis Baloyiannis, Jiannis Hajiioannou, Konstantinos Perivoliotis, George Tzovaras, General University Hospital of Larissa, Greece; Anna Fleck, Aine McGovern, Glasgow Royal Infirmary, UK; Victoria Gaunt, Gloucestershire Hospitals NHS Foundation Trust, UK; Laura Babb, Emily Bailey, Jay Darley, Ioan M Draghita, Alexander Hickman, Jason Kalloo, Akhil Kanzaria, Katy Madden, Wasim Nawaz, Ambreen Sadiq, Good Hope Hospital, UK; Rifa Cardoso, Margherita Faulkner, William Hurst, Ellen James, Aimee Leadbetter, Jordan Mayer, Tanya Robinson, Emma Stratton, Miriam Thake, Hannah Thould, Hannah Watson, Great Western Hospital, UK; Ravindra Belgamwar, Corrina Bentley, Harplands Hospital, UK; Sarah Freshwater, Health Education West Midlands, UK; Sergio DV Ruiz, Nuria M Sanz, Milagros Carrasco Prats, Pedro VF Fernández, Clara G Francés, Esther M Manuel, Miguel R Marin, Pedro L Morales, Patricia P Pérez, María V Soriano, Ismael Mora-Guzmán, Hospital General Reina Sofía, Spain; Ismael Mora-Guzmán, Hospital Santa Bárbara, Spain; Melanie Dani, Imperial College Healthcare NHS Trust, UK; Fabio Barra, Antonella Ferraiolo, Simone Ferrero, Claudio Gustavino, Chiara Kratochwila, IRCCS Ospedale Policlinico San Martino, Italy; Eric W Etchill, Alodia Gabre-Kidan, Joshua H Gray, Elliott R Haut, Harsha Malapati, Sarah F Rapaport, Kent A Stevens, Dominique Vervoort, Johns Hopkins Hospital, USA; Mohammed A Azab, King Abdullah Medical City Specialist Hospital, Saudi Arabia; Catherine Bryant, Hannah Cheney-Lowe, Catrin Cox, Andrew Crowe, Gordon Dick, Sarah Evans, Patrick CP Hogan, Kar Yee Law, Alexandra Richardson, Fabio Speranza, Kathryn Toppley, Julie Whitney, Eirene Yeung, King’s College Hospital, UK; Mary Ni Lochlainn, Claire Steves, King’s College London, UK; Alexandros Charalabopoulos, Spyridon Davakis, Amalia Karapanou, Theodore Liakakos, Eustratia Mpaili, Maria Mpoura, Michail A Sampanis, Nikolaos V Sipsas, Laiko University Hospital, Greece; Lucy Beishon, Elinor Burn, Parveen Doddamani, Victoria Haunton, Shahriar Kabir, Hannah Shaw, Chloe Warner, Leicester Royal Infirmary, UK; Chee Soo, Maidstone and Tunbridge Wells NHS Trust, UK; Yasmin K NasrEldin, Minia University Hospital, Egypt; Nourhan AA Ghannam, Minya General Hospital, Egypt; Isobel Sleeman, NHS Grampian, UK; Ravindra Belgamwar, Corrina Bentley, North Staffordshire Combined Healthcare NHS Trust, UK; Ali Ali, Sylvia Amini, James Belcher, Marie Giles, Hayley Jarvis, Nathan Jenko, Suvira Madan, Alexander Noar, Favour Nwolu, Jessica Parkin, Lauren C Passby, Jarita Sivam, Michael Surtees, Joanne Wagland, Ruth West, David Williams, Northern General Hospital, UK; Avinash Aujayeb, Lindsey Dew, Catherine Dotchin, James M Dundas, Elinor Edwards, Georgia F Gilbert, Karl Jackson, Sarah H Manning, Dominic Maxfield, Nicholas Moss, Declan C Murphy, Ellen Tullo, Sarah H Welsh, Northumbria NHS Hospital Trust, UK; Tahir Masud, Nottingham University Hospitals NHS Trust, UK; Mustafa Alsahab, Oxford University Hospitals NHS Trust, UK; Antonio Buondonno, Enrico Pinotti, Policlinico San Pietro, Italy; Francesco Alessandri, Gioia Brachini, Giancarlo Ceccarelli, Flavia Ciccarone, Pierfranco M Cicerchia, Bruno Cirillo, Giorgio De Toma, Giulia Duranti, Enrico Fiori, Giovanni B Fonsi, Pierfrancesco Lapolla, Simona Meneghini, Andrea Mingoli, Francesco Pugliese, Paolo Sapienza, Luigi Simonelli, Martina Zambon, Policlinico Umberto I, Sapienza University of Rome, Italy; Caterina Cattel, Laurenny Guzman, Princess Royal Hospital, King’s College Hospital Trust, Surrey, UK; Hannah Dowell, Aina Ibukunoluwakitan, Fawsiya Mohamed, Claire Spice, Amanda Stafford, Queen Alexandra Hospital, Portsmouth, UK; Jolene Atia, Catherine Atkin, Hannah Currie, Felicity Evison, Heena Khiroya, Zeinab Majid, Maria Qurashi, Queen Elizabeth Hospital Birmingham, UK; Siobhan Coulter, Claire McDonald, Georgina Muir, Catherine O’Mahony, Caroline Tait, Queen Elizabeth Hospital Gateshead, UK; Rowan Davies, Katie Honney, Laura Winter, Queen Elizabeth Hospital King’s Lynn, UK; Olubayode Adewole, Queen’s Hospital Romford, UK; Amir Abdelmalak, Mohammed Ahmad, Muhammed H Ansari, Kingsley Appiah, Rajesh Dwivedi, Hope Elrick, Hedra Ghobrial, Rosie Jackson, Sophie Jeffs, Sasha Jeyakumar, Eleanor Lunt, Bushra Muzammil, Sylvia Pytraczyk, Jonathan Sheldrake, Jennifer Smith, Hannah Tobiss, Mark Vettasseri, Ruth H Willott, Hein Zaw, Queens Medical Centre, Nottingham, UK; Katherine Patterson, Queen’s University Belfast, UK; Moulinath Bannerjee, Jean Cummings, Barbara Hart, Tom Maughan, Royal Bolton Hospital, UK; Clare Baguneid, Gabrielle Budd, Lizzie Moriarty, Omoteniola Odutola, Hannah Street, Royal Derby Hospital, UK; Alexis Carr, Royal Devon and Exeter Hospital, UK; Jennifer Pigott, Royal Free London NHS Foundation Trust, UK; Sarah Baldwin, Hannah Bashir, Jake Gibbon, Amy Gray, Grace Lewis, Christina Page, Rosanna Varden, Royal Victoria Infirmary, UK; Anthony Grubb, Elizabeth Holmes, Harjinder Kainth, Natalie McNeela, Lara Reilly, Abigail Reynolds, Mark Whitsey, Royal Wolverhampton NHS Trust, UK; Mertcan Akcay, Yeşim Akdeniz, Emrah Akin, Fatih Altintoprak, Zülfü Bayhan, Recayi Capoglu, Hakan Demir, Necattin Firat, Emre Gonullu, Tarık Harmantepe, Baris Mantoglu, Ali Muhtaroglu, Merve Yigit, Yasin A Yildiz, Sakarya Faculty of Medicine, Turkey; Lobna Al-Sodani, Nicole Burden, Evelyn Charsley, Thomas Kneen, Angeline Price, Emma Swinnerton, Salford Royal Hospital, UK; Yen Nee J Bo, Hayley R Boden, Reem Bulla, Alison Eastaugh, Helena Lee, Asma Khan, Mohammed Mubin, Amelia Roberts, Anthony Umeadi, Stephanie Wallis, Megan Williamson, Yu Lelt Win, Sandwell General Hospital, UK; Eltayeb A Ahmed, Abdulmoiz Aljafari, Abdulmalek Aljafari, Abdulkader Mohammad, Sharq Alneel Hospital, Sudan; Manpreet Badh, Amy Birchenough, Nick Coulthard, Alice Devaney, Ratnam Gandhi, Katharine Hood, Samuel North, Martha Pinkney, Ellie Shaw, Elisha Whelan, Solihull Hospital, UK; Adam Seed, Southport and Ormskirk Hospital NHS Trust, UK; Gurinder Dogra, Claire Morris, Rebecca Wright, South Tyneside District Hospital, UK; Stephen Lim, Lia Orlando, Harnish Patel, Prabhleen Puri, Sing Yang Sim, Southampton General Hospital, UK; Carolyn Akladious, Gitanjali Amaratungaz, Taha Amir, Cheran Anandarajah, Rachael Anders, Sally Aziz, Anna Barnard, Monica Bawor, Laura Bremner, Hannah Bridgwater, Hejab Butt, Andra Caracostea, Theodore Chevallier, Victoria Comerford, Jack Cullen, Niamh Cunningham, Daniel Curley, Madeleine Daly, Nikhita Dattani, Benyamin Deldar, Arjun Desai, Nirali Desai, Jugdeep Dhesi, Maria Dias, Hannah C Dooley, Samiullah Dost, Hiren Dusara, Alexander Emery, Cassandra Fairhead, Antia Fernandez, Gracie Fisk, Madeleine Garner, Hannah Gerretsen, Andrew Ghobrial, Zaynub Ghufoor, Deirdre Green, Charlotte Greene, Karla Griffith, Ayushi Gupta, Patrick Harrison, Aidan Haslam, Torben Heinsohn, Lindsay Hennah, Abigail Hobill, Katherine Hopkinson, Lara Howells, Nicole Hrouda, Irem Ishlek, Rishi Iyer, Nuha Kardaman, Mairead Kelly, Nicola I Kelly, Hesham Khalid, Muhammad S Khan, Haris Khan, Matthew King, Li Kok, Aneliya Kuzeva, Rebecca Lau, Gabriel Lee, Gavriella Levinson, Danielle Lis, Baguiasri Mandane, Jamie Mawhinney, Henry Maynard, Sophie Mclachlan, Michelle Metcalf, John Millwood-Hargrave, Kelvin Miu, Aaliya Mohammed, Hamilton Morrin, Stephanie Mulhern, Daniel Muller, Varun Nadkarni, Hanna Nguyen, Alice O’Docherty, Sinead O’Dwyer, Marc Osterdahl, Ismini Panayotidis, Shefali Patel, Rose Penfold, Rupini Perinpanathan, Dina Radenkovic, Thurkka Rajeswaran, Tahmina Razzak, Emily Ross-Skinner, Hazel Sanghvi, Ross Sayers, Luca Scott, Sri Sivarajan, Katharine Stambollouian, Jack Stewart, Amybel Taylor, Hrisheekesh Vaidya, Vittoria Vergani, Madiha Virk, Vaishali Vyas, Eleanor Watkins, Catherine Wilcock, Mettha Wimalasundera, Stephanie Worrall, Natalie Yeo, Humza Yusuf, St Thomas’ Hospital, UK; Adam H Dyer, Cliona Ni Cheallaigh, Liam Townsend, St. James’s Hospital, Ireland; Jocelyn Amer, Emily Lyon, Michael Sen, Sunderland Royal Hospital, UK; Mohammed Al-Sadawi, Adam Budzikoski, Ishmam Ibtida, Yusra Qaiser, SUNY Downstate Brooklyn, USA; Mohammad T Azam, Asad J Choudhry, William Marx, SUNY Upstate University Hospital, USA; Ahmad Bouhuwaish, Ahmed SA Taher, Tobruk Medical Center, Libya; Nikolaos Georgiou, Jade Man, Paul Reynolds, Benjaman To, Tunbridge Wells Hospital, UK; Fatma D Collins, Sharon Budd, Ellanna Griffin, Yue Guan, Deevia Hanji, Lily Lowes, Awolkhier Mohammedseid-Nurhussien, Farhana Moomo, Olebu Ogochukwu, Katie Thin, University Hospitals Coventry and Warwickshire NHS Trust, UK; Elinor Burn, University Hospitals of Leicester NHS Trust; Terry Hughes, Thomas A Jackson, Laura Magill, Lauren McCluskey, Hannah Moorey, Kelvin Okoth, Rita Perry, Michala Petitt, Thomas Pinkney, Daisy Wilson, University of Birmingham; Grace ME Pearson, University of Bristol, UK; Christopher N Osuafor, Kelli Torsney, University of Cambridge, UK; David Strain, Jane Masoli, University of Exeter, UK; Jenni Burton, Terence Quinn, University of Glasgow, UK; Lucy Beishon, University of Leicester, UK; Joanne Taylor, University of Manchester, UK; Adam Gordon, University of Nottingham, UK; Gilda De Paola, Gaetano Gallo, Giuseppe Sammarco, Giuseppina Vescio, University ‘Magna Graecia’ of Catanzaro, Italy; Shivam Pancholi, University of Nicosia, Cyprus; Natalie Cox, University of Southampton, UK; Rajni Lal, Western Sydney Local Health District, Australia; Rand A Hussein, Zafaraniyah General Hospital, Iraq.

