## Supplementary tables for "Development and external validation of prognostic models for COVID-19 to support risk stratification in secondary care"

**Supplementary Table 1. Area under the receiver operating characteristic curve (AUROC) for each of the models assessed in the derivation dataset (University Hospitals Birmingham) for predicting death within 28 days of hospital admission**

| **Model** | **Continuous data** | **Missing continuous data** | **Candidate predictors included** | **AUROC (95% CI)** |
| --- | --- | --- | --- | --- |
| Logistic | Categorical | Imputed | All | 0.744 (0.709-0.780) |
| LASSO | Categorical | Imputed | All | 0.751 (0.716-0.786) |
| GBM | Categorical | Imputed | All | 0.756 (0.721-0.790) |
| Logistic | Categorical | Imputed | Stepwise selection | 0.778 (0.741-0.815) |
| Logistic | Categorical | Imputed | All plus comorbidities | 0.734 (0.697-0.771) |
| Logistic | Categorical | Imputed | Demographic only | 0.709 (0.674-0.744) |
| Logistic | Categorical | Missing category | All | 0.766 (0.725-0.808) |
| Logistic | Categorical | Missing category | Demographic only | 0.705 (0.670-0.740) |
| Logistic | Categorical | Missing category | Stepwise selection | 0.805 (0.777-0.834) |
| LASSO | Categorical | Missing category | All | 0.781 (0.745-0.817) |
| GBM | Categorical | Missing category | All | 0.776 (0.739-0.813) |
| Logistic | Continuous | Imputed | All | 0.765 (0.724-0.806) |
| Logistic | Continuous | Imputed | All with splines* | 0.719 (0.633-0.805) |
| LASSO | Continuous | Imputed | All | 0.777 (0.745-0.808) |
| LASSO | Continuous | Imputed | All with splines* | 0.782 (0.752-0.812) |
| GBM | Continuous | Imputed | All | 0.773 (0.741-0.804) |
| GBM | Continuous | Imputed | All with splines* | 0.773 (0.739-0.808) |
| Logistic | Continuous | Imputed | Stepwise selection | 0.794 (0.764-0.823) |
| Logistic | Continuous | Imputed | Stepwise selection with splines* | 0.791 (0.759-0.823) |

All = all demographic, symptoms, vital signs/physiological measures, and laboratory test result candidate predictors. *Splines were used to model continuous variables where non-liner associations with the outcome were observed. LASSO = least absolute shrinkage and selection operator; GBM = gradient boosting machine.

**Supplementary Table 2. Calibration slopes and intercepts for model performance in the University Hospitals Birmingham (UHB) derivation datasets (full (UHB) and reduced (UHB-R) datasets) and in the CovidCollab external validation dataset**

| **Dataset** | **Outcome** | **Calibration** | |
| --- | --- | --- | --- |
|  |  | **Slope** | **Intercept** |
| **Model development*** |  |  |  |
| UHB | Mortality | 0.78 (0.66-0.91) | 0.06 (0.00-0.13) |
| UHB | ITU admission | 0.89 (0.77-1.02) | 0.02 (-0.05-0.09) |
| UHB-R† | Mortality | 0.86 (0.82-0.90) | 0.04 (0.02-0.06) |
| UHB-R† | ITU admission | 0.94 (0.84-1.03) | 0.01 (-0.05-0.06) |
| **External validation of new model** | | |  |
| CovidCollab | Mortality | 0.88 (0.80-0.96) | 0.03 (-0.01-0.07) |
| CovidCollab | ITU admission | 0.93 (0.84-1.02) | 0.01 (-0.04-0.06) |

*Models derived using logistic regression with stepwise selection of candidate predictors and categorisation of continuous variables into clinically meaningful categories (after imputing missing data).

†Not all variables included in the full University Hospital Birmingham (UHB) model were available in the CovidCollab dataset. Therefore, revised (reduced) models were developed in UHB data using a subset of the candidate predictors common to both the UHB and CovidCollab datasets (UHB-R), and these were then externally validated in the CovidCollab dataset.

**Supplementary Table 3. Logistic regression model coefficients for mortality within 28 days of admission in the derivation dataset (University Hospitals Birmingham)**

| **Predictor** | **Coefficient** |
| --- | --- |
| Intercept | -16.312 |
| Age category (years), n (%) |  |
| <30 | Ref |
| 30-39 | 13.508 |
| 40-49 | 12.722 |
| 50-59 | 14.158 |
| 60-69 | 14.350 |
| 70-79 | 15.160 |
| 80-89 | 15.570 |
| ≥90 | 15.864 |
| Breathlessness | 0.304 |
| Sputum | 0.442 |
| Temperature (Celsius), n (%) |  |
| <37.8 | Ref |
| ≥37.8 | 0.339 |
| Systolic blood pressure (mm Hg), n (%) |  |
| <140 | Ref |
| ≥140 | -0.360 |
| Respirations (per minute), n (%) |  |
| <20 | Ref |
| ≥20 | 0.358 |
| Oxygen saturation (%), n (%) |  |
| <80 | Ref |
| 80-88 | 0.319 |
| 89-93 | -0.808 |
| ≥94 | -1.135 |
| Portable oxygen concentrator fraction of inspired oxygen (%), n (%) |  |
| ≤0.28 | Ref |
| 0.28-0.49 | 0.518 |
| ≥0.5 | 0.627 |
| pH, n (%) |  |
| <7.30 | Ref |
| 7.30-7.34 | -1.017 |
| 7.35-7.44 | -0.486 |
| ≥7.45 | -0.406 |
| White blood cell count (10^9^/l), n (%) |  |
| <3.9 | Ref |
| 3.9-10.8 | 0.915 |
| ≥10.9 | 0.999 |
| Platelets (10^9^/l), n (%) |  |
| <150 | Ref |
| 150-399 | -0.507 |
| ≥400 | -0.792 |
| C-reactive protein (mg/l), n (%) |  |
| <10 | Ref |
| 10-99 | 0.530 |
| ≥100 | 1.014 |
| Glucose (mmol/l), n (%) |  |
| <7.8 | Ref |
| 7.8-8.4 | 0.571 |
| ≥8.5 | 0.426 |
| Alkaline phosphatase (U/l), n (%) |  |
| <130 | Ref |
| ≥130 | 0.389 |
| Urea (mmol/l), n (%) |  |
| <7.8 | Ref |
| ≥7.8 | 0.376 |
| Corrected calcium (mmol/l), n (%) |  |
| <2.2 | Ref |
| 2.2-2.5 | -0.695 |
| ≥2.6 | 0.503 |
| Eosinophils (10^9^/l), n (%) |  |
| ≤0.4 | Ref |
| >0.4 | -1.408 |
| Frailty score, n (%) |  |
| 1-3 |  |
| 4-6 | 0.170 |
| 7-9 | 0.722 |

(Coefficients derived after a single imputation of missing data.)

**Supplementary Table 4. Area under the receiver operating characteristic curve (AUROC) for each of the models assessed in the derivation dataset (University Hospitals Birmingham) for predicting intensive therapy unit admission within 28 days of hospital admission**

| **Model** | **Continuous data** | **Missing continuous data** | **Candidate predictors included** | **AUROC (95% CI)** |
| --- | --- | --- | --- | --- |
| Logistic | Categorical | Imputed | All | 0.846 (0.813-0.880) |
| LASSO | Categorical | Imputed | All | 0.872 (0.842-0.901) |
| GBM | Categorical | Imputed | All | 0.875 (0.843-0.906) |
| Logistic | Categorical | Imputed | Stepwise selection | 0.892 (0.865-0.920) |
| Logistic | Categorical | Imputed | All plus comorbidities | 0.870 (0.836-0.904) |
| Logistic | Categorical | Imputed | Demographic only | 0.763 (0.728-0.798) |
| Logistic | Categorical | Missing category | All | 0.911 (0.867-0.954) |
| Logistic | Categorical | Missing category | Demographic only | 0.767 (0.734-0.800) |
| Logistic | Categorical | Missing category | Stepwise selection | 0.955 (0.933-0.978) |
| LASSO | Categorical | Missing category | All | 0.962 (0.947-0.976) |
| GBM | Categorical | Missing category | All | 0.954 (0.938-0.970) |
| Logistic | Continuous | Imputed | All | 0.869 (0.833-0.905) |
| Logistic | Continuous | Imputed | All with splines* | 0.891 (0.864-0.919) |
| LASSO | Continuous | Imputed | All | 0.885 (0.857-0.914) |
| LASSO | Continuous | Imputed | All with splines* | 0.905 (0.88-00.931) |
| GBM | Continuous | Imputed | All | 0.897 (0.871-0.924) |
| GBM | Continuous | Imputed | All with splines* | 0.899 (0.870-0.928) |
| Logistic | Continuous | Imputed | Stepwise selection | 0.894 (0.867-0.921) |
| Logistic | Continuous | Imputed | Stepwise selection with splines* | 0.908 (0.883-0.934) |

All = all demographic, symptoms, vital signs/physiological measures, and laboratory test result candidate predictors. *Splines were used to model continuous variables where non-liner associations with the outcome were observed. LASSO = least absolute shrinkage and selection operator; GBM = gradient boosting machine.

**Supplementary Table 5. Logistic regression model coefficients for intensive therapy unit admission within 28 days of hospital admission in the derivation dataset (University Hospitals Birmingham)**

| **Predictor** | **Coefficient** |
| --- | --- |
| Intercept | -1.394 |
| Age category (years), n (%) |  |
| <30 | Ref |
| 30-39 | 0.221 |
| 40-49 | 0.637 |
| 50-59 | 1.046 |
| 60-69 | 0.296 |
| 70-79 | -0.282 |
| 80-89 | -2.240 |
| ≥90 | -15.912 |
| Sex (male) | 0.791 |
| Fever | 0.429 |
| New onset diarrhoea or vomiting | -0.808 |
| Temperature (Celsius), n (%) |  |
| <37.8 | Ref |
| ≥37.8 | 0.605 |
| Heart rate category (beats per minute), n (%) |  |
| <80 | Ref |
| 80-99 | -0.572 |
| ≥100 | -0.818 |
| Respirations (per minute), n (%) |  |
| <20 | Ref |
| ≥20 | 0.541 |
| Portable oxygen concentrator fraction of inspired oxygen (%), n (%) |  |
| ≤0.28 | Ref |
| 0.28-0.49 | 1.206 |
| ≥0.5 | 1.766 |
| pH, n (%) |  |
| <7.30 | Ref |
| 7.30-7.34 | -1.912 |
| 7.35-7.44 | -1.740 |
| ≥7.45 | -1.243 |
| White blood cell count (10^9^/l), n (%) |  |
| <3.9 | Ref |
| 3.9-10.8 | 0.757 |
| ≥10.9 | 2.203 |
| C-reactive protein (mg/l), n (%) |  |
| <10 | Ref |
| 10-99 | 0.241 |
| ≥100 | 0.634 |
| Albumin (g/l), n (%) |  |
| <25 | Ref |
| 25-34 | -0.835 |
| ≥35 | -0.913 |
| Monocytes (10^9^/l), n (%) |  |
| <0.2 | Ref |
| 0.2-0.8 | 0.068 |
| >0.8 | -0.956 |
| eGFR (ml/min), n (%) |  |
| <30 (Stage 4 or above) | Ref |
| 30-59 (Stage 3) | 1.079 |
| 60-89 (Stage 2) | 0.524 |
| >90 (Normal or high) | 0.723 |
| Frailty score, n (%) |  |
| 1-3 | Ref |
| 4-6 | -0.519 |
| 7-9 | -1.929 |
| Glasgow Coma Scale score, n (%) |  |
| <15 | Ref |
| 15 | -1.182 |

(Coefficients derived after a single imputation of missing data.)

**Supplementary Table 6. Logistic regression model coefficients for the external validation model for mortality within 28 days of hospital admission (developed in University Hospitals Birmingham data and validated in CovidCollab)**

| **Predictor** | **Coefficient** |
| --- | --- |
| Intercept | -16.189 |
| Age category (years), n (%) |  |
| <30 | Ref |
| 30-39 | 13.569 |
| 40-49 | 12.853 |
| 50-59 | 14.234 |
| 60-69 | 14.451 |
| 70-79 | 15.180 |
| 80-89 | 15.461 |
| ≥90 | 15.803 |
| Temperature (Celsius), n (%) |  |
| <37.8 | Ref |
| ≥37.8 | 0.402 |
| Systolic blood pressure (mm Hg), n (%) |  |
| <140 | Ref |
| ≥140 | -0.361 |
| Respirations (per minute), n (%) |  |
| <20 | Ref |
| ≥20 | 0.412 |
| Oxygen saturation (%), n (%) |  |
| <80 | Ref |
| 80-88 | 0.452 |
| 89-93 | -0.564 |
| ≥94 | -1.045 |
| Portable oxygen concentrator fraction of inspired oxygen (%), n (%) |  |
| ≤0.28 | Ref |
| 0.28-0.49 | 0.562 |
| ≥0.5 | 0.632 |
| pH, n (%) |  |
| <7.30 | Ref |
| 7.30-7.34 | -0.945 |
| 7.35-7.44 | -0.535 |
| ≥7.45 | -0.352 |
| C-reactive protein (mg/l), n (%) |  |
| <10 | Ref |
| 10-99 | 0.607 |
| ≥100 | 1.110 |
| Urea (mmol/l), n (%) |  |
| <7.8 | Ref |
| ≥7.8 | 0.489 |
| Frailty score, n (%) |  |
| 1-3 | Ref |
| 4-6 | 0.166 |
| 7-9 | 0.659 |

(Coefficients derived after a single imputation of missing data.)

**Supplementary Table 7. Logistic regression model coefficients for the external validation model for intensive therapy unit admission within 28 days of hospital admission (developed in University Hospitals Birmingham data and validated in CovidCollab)**

| **Predictor** | **Coefficient** |
| --- | --- |
| Intercept | -2.155 |
| Age category (years), n (%) |  |
| <30 | Ref |
| 30-39 | 0.309 |
| 40-49 | 0.651 |
| 50-59 | 0.969 |
| 60-69 | 0.319 |
| 70-79 | -0.110 |
| 80-89 | -1.887 |
| ≥90 | -15.724 |
| Sex (male) | 0.683 |
| Fever | 0.448 |
| Respirations (per minute), n (%) |  |
| <20 | Ref |
| ≥20 | 0.433 |
| Portable oxygen concentrator fraction of inspired oxygen (%), n (%) |  |
| ≤0.28 | Ref |
| 0.28-0.49 | 1.180 |
| ≥0.5 | 1.816 |
| pH, n (%) |  |
| <7.30 | Ref |
| 7.30-7.34 | -1.776 |
| 7.35-7.44 | -1.925 |
| ≥7.45 | -1.388 |
| C-reactive protein (mg/l), n (%) |  |
| <10 | Ref |
| 10-99 | -0.018 |
| ≥100 | 0.612 |
| Neutrophil:lymphocyte ratio, n (%) |  |
| <2.21 | Ref |
| 2.21-4.82 | 0.747 |
| >4.82 | 1.143 |
| eGFR (ml/min), n (%) |  |
| <30 (Stage 4 or above) | Ref |
| 30-59 (Stage 3) | 0.936 |
| 60-89 (Stage 2) | 0.363 |
| >90 (Normal or high) | 0.606 |
| Frailty score, n (%) |  |
| 1-3 | Ref |
| 4-6 | -0.424 |
| 7-9 | -1.852 |
| Glasgow Coma Scale score, n (%) |  |
| <15 | Ref |
| 15 | -1.139 |

(Coefficients derived after a single imputation of missing data.)

**Supplementary table 8. Sensitivity analysis – complete case analysis and exploration of different numbers of missing candidate predictors: Area under the receiver operating characteristic curve (AUROC) in the University Hospitals Birmingham dataset for predicting mortality and intensive therapy unit admission within 28 days of hospital admission**

| **Number of missing predictor variables per patient** | **n** | **Outcome** | |
| --- | --- | --- | --- |
|  |  | **Death** | **ITU** |
| 0 | 224 | 0.690 (0.588-0.791) | 0.887 (0.825-0.949) |
| 0-1 | 471 | 0.767 (0.715-0.819) | 0.885 (0.843-0.927) |
| 0-2 | 564 | 0.761 (0.711-0.811) | 0.880 (0.848-0.912) |
| 0-5 | 643 | 0.753 (0.709-0.797) | 0.890 (0.860-0.921) |
| 0-10 | 911 | 0.770 (0.735-0.805) | 0.883 (0.855-0.912) |

**Supplementary table 9. Sensitivity analysis – stratification by gender: Area under the receiver operating characteristic curve (AUROC) in the University Hospitals Birmingham dataset for predicting mortality and intensive therapy unit admission within 28 days of hospital admission**

| **Model** | | **Outcome** | |
| --- | --- | --- | --- |
| **Derivation/Train** | **Test** | **Death** | **ITU** |
| Fitted on all patients | Male | 0.766 (0.700-0.831) | 0.893 (0.823-0.963) |
| Fitted on all patients | Female | 0.788 (0.729-0.847) | 0.893 (0.851-0.935) |
| Fitted on males | Male | 0.738 (0.678-0.798) | 0.837 (0.770-0.905) |
| Fitted on females | Female | 0.779 (0.726-0.832) | 0.884 (0.842-0.926) |

**Supplementary figure legends:**

Supplementary Figure 1. Exploration of non-linear relationships between continuous predictors and 28-day mortality using general additive models (GAM); splines (continuous smoothed predictors) were included in the models where a non-linear relationship was observed.

Supplementary Figure 2. Exploration of non-linear relationships between continuous predictors and intensive therapy unit admission using general additive models (GAM); splines (continuous smoothed predictors) were included in the models where a non-linear relationship was observed.

Supplementary Figure 3. Forest plot of logistic regression coefficients for days 0-7 from admission for mortality outcome.
