## Supplementary figures and images for "Development and external validation of prognostic models for COVID-19 to support risk stratification in secondary care"

### Supplementary figure 1

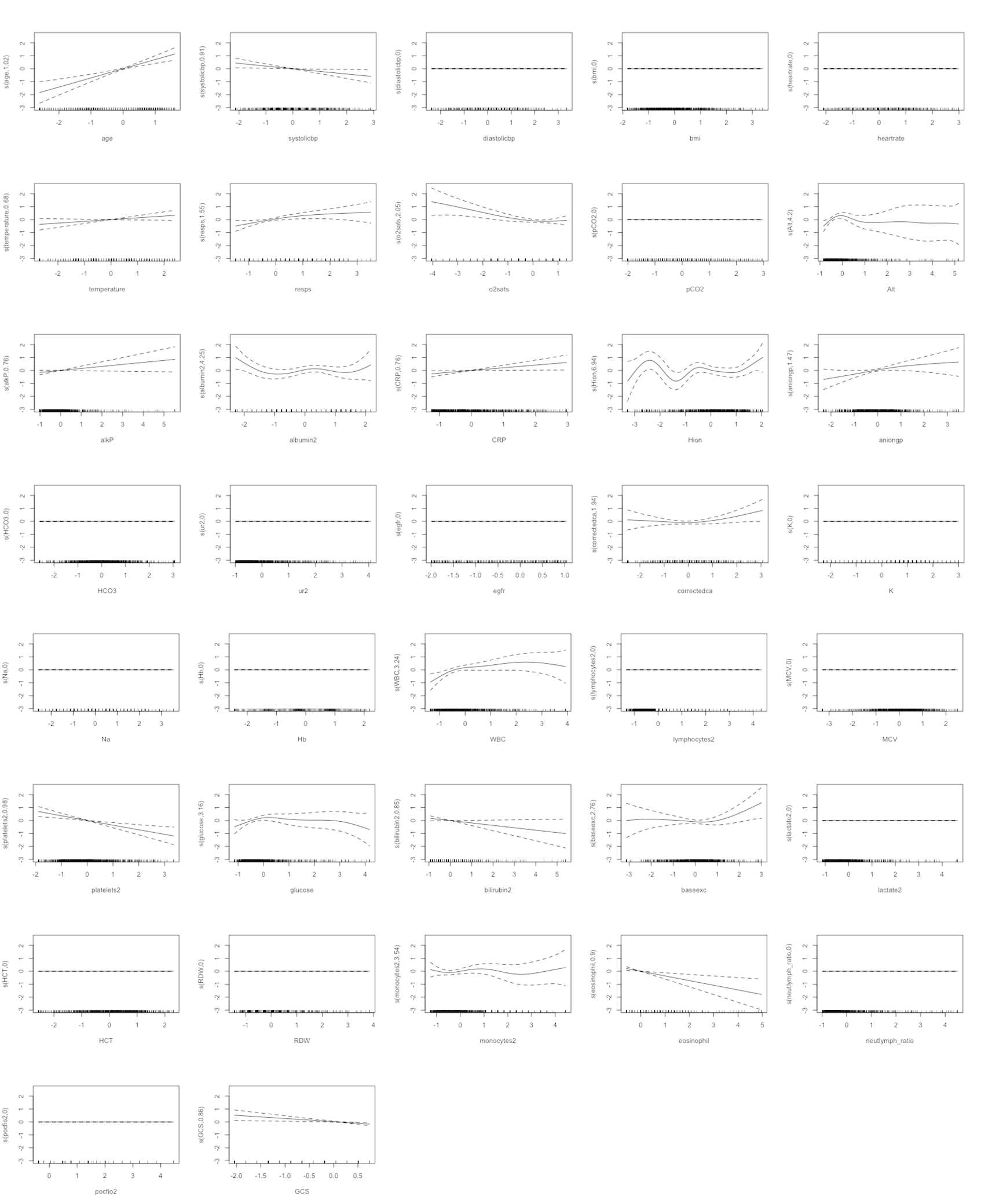

### Supplementary figure 2

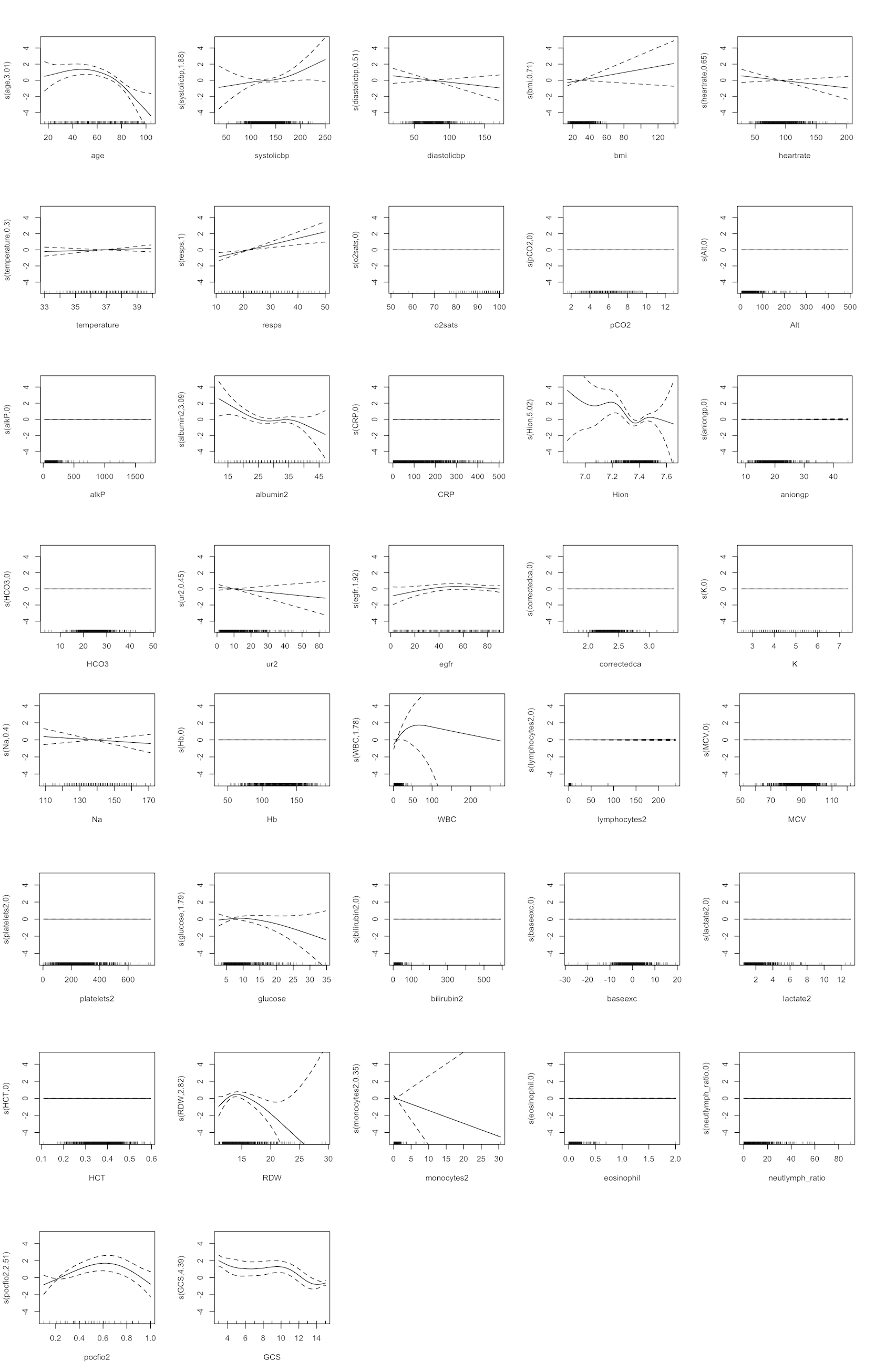

### Supplementary figure 3

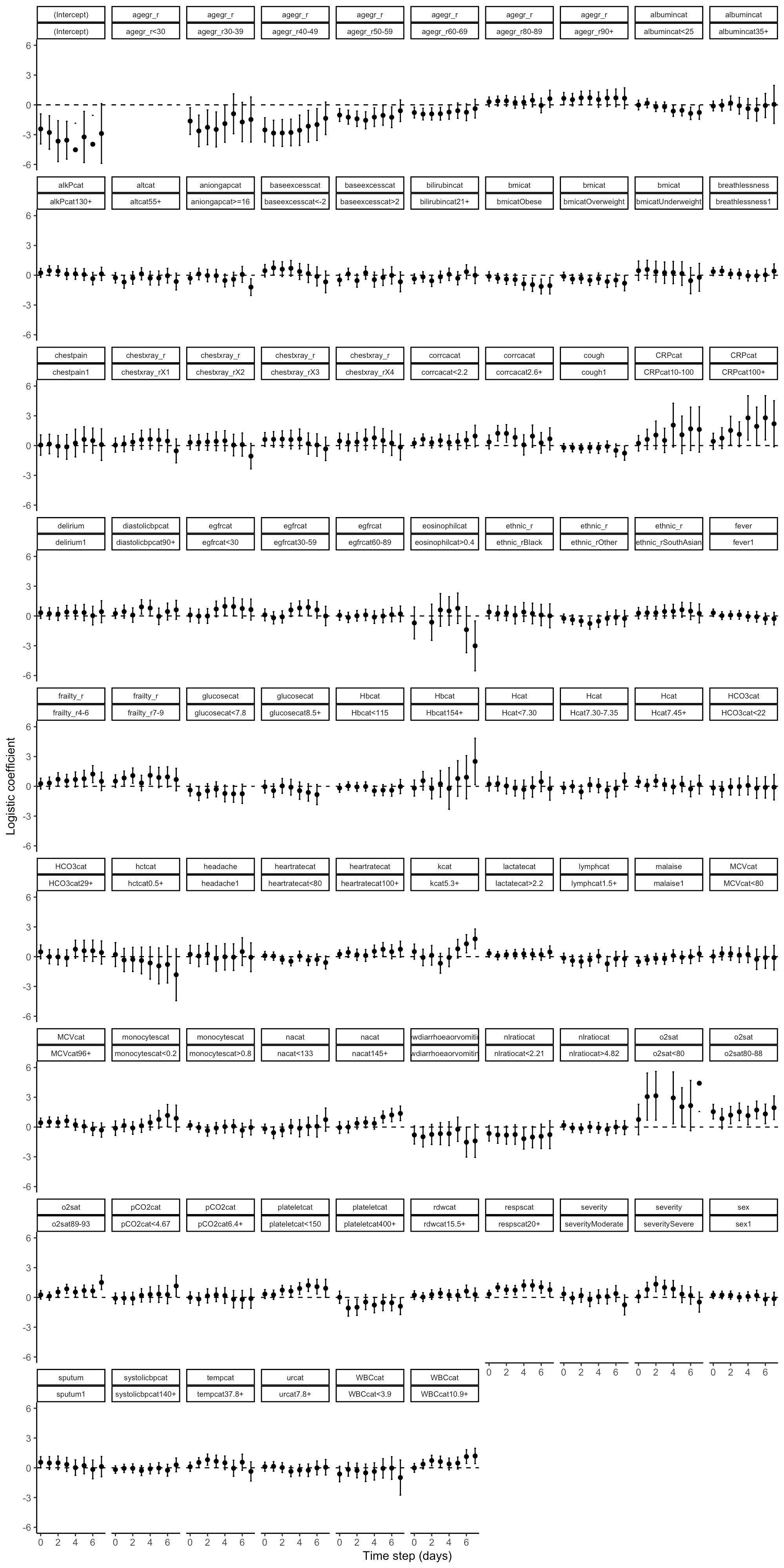
